# Next-generation Serology by Mass Spectrometry: Readout of the SARS-CoV-2 Antibody Repertoire

**DOI:** 10.1101/2021.07.06.21259226

**Authors:** Rafael D. Melani, Benjamin J. Des Soye, Jared O. Kafader, Eleonora Forte, Michael Hollas, Voislav Blagojevic, Fernanda Negrão, John P. McGee, Bryon Drown, Cameron Lloyd-Jones, Henrique S. Seckler, Jeannie M. Camarillo, Philip D. Compton, Richard D. LeDuc, Bryan Early, Ryan T. Fellers, Byoung-Kyu Cho, Basil Baby Mattamana, Young Ah Goo, Paul M. Thomas, Michelle K. Ash, Pavan P. Bhimalli, Lena Al-Harthi, Beverly E. Sha, Jeffrey R. Schneider, Neil L. Kelleher

**Affiliations:** Departments of Molecular Biosciences, Chemistry, Chemical and Biological Engineering, and the Feinberg School of Medicine, Northwestern University, Evanston, IL, USA; Proteomics Center of Excellence, Evanston, IL, USA; Department of Surgery, Feinberg School of Medicine, Northwestern University, Chicago, IL, USA; Integrated Protein Technologies, Evanston, IL, USA; Department of Microbial Pathogens and Immunity, Rush University Medical Center, Chicago, IL, USA; Division of Infectious Diseases, Rush University Medical Center, Chicago, IL, USA

**Author notes:** Correspondence to: Neil L Kelleher, 2145 Sheridan Rd, Evanston, IL 60208. These authors contributed equally to this work (Rafael D. Melani, Benjamin J. Des Soye, Jared O. Kafader, Eleonora Forte).

## Abstract

Methods of antibody detection are used to assess exposure or immunity to a pathogen. Here, we present <u>Ig-MS</u>, a novel serological readout that captures the immunoglobulin (Ig) repertoire at molecular resolution, including entire variable regions in Ig light and heavy chains. Ig-MS uses recent advances in protein mass spectrometry (MS) for multi-parametric readout of antibodies, with new metrics like Ion Titer (IT) and Degree of Clonality (DoC) capturing the heterogeneity and relative abundance of individual clones without sequencing of B cells. We apply Ig-MS to plasma from subjects with severe & mild COVID-19, using the receptor-binding domain (RBD) of the spike protein of SARS-CoV-2 as the bait for antibody capture. Importantly, we report a new data type for human serology, with compatibility to any recombinant antigen to gauge our immune responses to vaccination, pathogens, or autoimmune disorders.

## Main

Since late 2019, severe acute respiratory syndrome coronavirus 2 (SARS-CoV-2) and its associated disease COVID-19^1^ have been at the center of a global pandemic affecting more than 150 million people, especially immunocompromised individuals, the elderly, and individuals with pre-existing conditions^2^. This has resulted in >3 million deaths worldwide, including ∼600,000 in the USA where COVID-19 became a leading cause of death^3, 4^. Unfortunately, the global death toll is still rising due to delays in vaccination and the emergence of more transmissible variants of SARS-CoV-2, against which the available vaccines and antibody-based therapeutics may be less effective^4^.

Most COVID-19 patients develop antibodies against SARS-CoV-2 within a few weeks after infection^5, 6^, commonly recognizing targets such as viral envelope, nucleocapsid, and spike proteins. In particular, the spike protein receptor-binding domain (RBD) is a robust immunogenic target that has been the focus of many antibody-based diagnostics and therapeutics against SARS-CoV-2^7^.

Generally, standard serology tests (*i*.*e*., ELISA and lateral flow assays) can reliably detect whether a patient possesses antibodies against a target antigen, but they do not capture the clonality and makeup of an entire immunoglobulin (Ig) repertoire. A more precise assessment of immune status relative to SARS-CoV-2 at single clone resolution would significantly improve serological testing capabilities. A proteomics approach to the antibody repertoire has been challenging, though digestion-based approaches combined with Ig-seq have been evaluated and attempted^8, 9^. Several studies have used a low-resolution view of the IgG landscape using peptide-based analyses of IgGs after tryptic digestion^10, 11^, including one conducted in the COVID-19 context^12, 13^.

Intact proteins have many proteoforms^14^, and in the case of antibody heterogeneity this could be captured if there was a molecular readout with enough resolution and specificity. Past efforts have been able to detect monogammapathies (*e*.*g*., B cell cancers like multiple myeloma^15^), but a direct readout of Ig repertoire at high resolution is not currently possible.

Addressing this, we established Ig-MS, which isolates antibodies against a specific target from the plasma of patients and controllably breaks them down into their heavy and light chains (HC and LC). Once created, HC and LC fragments are analyzed by individual ion mass spectrometry (I^2^MS), a breakthrough approach that generates mass distributions for extremely heterogeneous protein samples using ∼500 fold more dilute samples than classical protein MS analysis^16^. Using one of the two workflows (**Fig. 1a**), Ig-MS provides compositional profiles for Ig repertoires, their degree of clonality, and titers for an initial cohort of COVID-19 subjects. With increased resolution for molecular serology, Ig-MS could provide high-value clinical correlates of viral neutralization, the presence, extent, and course of the disease, and/or assess the degree of protection after COVID-19 vaccination.

**Fig. 1.**
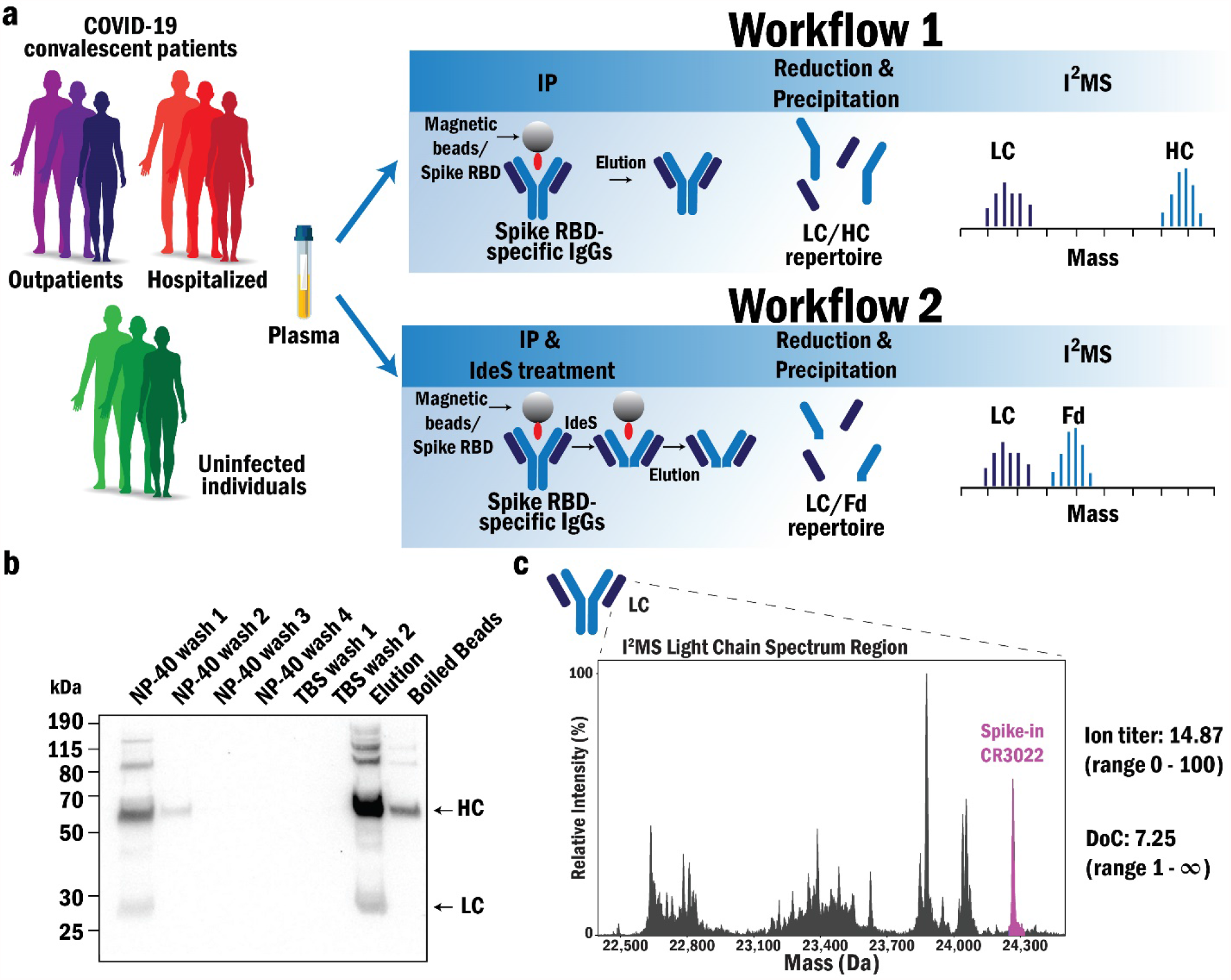
Ig-MS, a new platform for COVID-19 serology. (a) Overview of sample preparation and readout. Antibodies against the RBD domain of the SARS-CoV-2 spike protein are enriched from plasma of COVID-19 patients and controls using magnetic beads. Before elution, Spike RBD-specific Igs are either eluted (workflow 1) or treated with the IdeS enzyme to remove the Fc domain of the HC (workflow 2). The eluates are then reduced to obtain LC and HC (workflow 1) or LC and Fd (workflow 2), which are analyzed by individual ion mass spectrometry (I^2^MS). (b) Western blot showing the enrichment of Ig targeting SARS-CoV-2 Spike RBD from CS1 using workflow 1. (c) Example data set and metrics obtained from Ig-MS of the plasma from a COVID-19 convalescent patient (COVID-19_3) (only LC is shown).

## Results

### Benchmarking the Ig-MS Assay

The RBD of the spike protein from SARS-CoV-2 (Wuhan strain) was expressed, purified from HEK293 cells, and characterized by both bottom-up proteomics (100% sequence coverage) and western blot (**Supplementary Fig. 1**). Purified RBD was covalently attached to magnetic beads and used as bait for binding and enriching antibodies against SARS-CoV-2-Spike-RBD (Ig-RBD) from the plasma of COVID-19 convalescent individuals (**Fig. 1a**). One hundred nanograms of a standard mAb CR3022 was used as an internal standard in all Ig-MS (see **Online Methods**).

Proof-of-concept experiments used samples derived from a single COVID-19 convalescent individual featuring a high ELISA titer of Ig-RBD, hereafter referred to as commercial sample 1 (CS1). Enrichments from plasma derived from CS1 were performed to optimize parameters such as covalent bead-loading chemistry, the wash, and elution buffer conditions to increase capture and suppress non-specific binding (see **Online Methods**). As shown in **Fig. 1b**, the optimized pull-down protocol for 100 µL of plasma efficiently captured Ig-RBD, which was isolated with >90% yield in the optimized elution conditions. Pull-downs from CS1 were used to profile the antibody isotypes (IgA, IgD, IgE, and IgM) and IgG subclasses (IgG1, IgG2, IgG3, and IgG4) being enriched. A specific western blot revealed that IgG1, IgG3, and IgM were the primary isotypes isolated (**Supplementary Fig. 2**), similar to what has been reported in the literature^17^.

Following antibody enrichment, two distinct workflows were developed to prepare isolated Ig-RBD for Ig-MS analysis (**Fig. 1a**). In **workflow 1**, Ig-RBDs were eluted intact, combined with 100 ng of standard mAb CR3022, and fully reduced/denatured in the presence of a chaotropic agent to liberate HC (48-55 kDa) and LC (22-25 kDa) proteoforms. In **workflow 2**, patient Ig-RBD were digested with IdeS protease while still attached to the beads, generating F(ab’)_2_ and Fc. In **workflow 2**, the protease and Fc species (containing HC glycosylation) are washed away. Eluted F(ab’) _2_ were denatured and reduced to yield the LC and the Fd domain (∼25-28 kDa), which contains the N-terminal 220 amino acids of the HC and therefore captures the composition of all three Complementarity-Determining Regions (CDRs). Before the I^2^MS analysis step of Ig-MS, 100 ng of digested and reduced mAb CR3022 were spiked into the sample.

To benchmark Ig-MS, we first analyzed mAb CR3022 and annotated the LC and HC proteoforms (**Supplementary Fig. 3**). Subsequently, we estimated the Ig-MS Limit of Detection (LOD) for the HC of NIST mAb (with glycosylation) at 100 nM, whereas the LOD for the LC was well below 10 nM (**Supplementary Fig. 4**).

Next, we performed Ig-MS using workflow 1 on CS1, and **Supplementary Fig. 5** shows the spectrum obtained for the LC region. Like in **Fig. 1c**, the highlighted peak at ∼24,268 Da represents the mAb CR3022 standard used to calculate Ig-MS metrics, as outlined in **Equations 1-3** in the **Online Methods**. The other peaks with lower mass than the standard LC, ranging from ∼22 to 24 kDa, are distinct LC proteoforms originating from B cell clones with different or isobaric CDR sequences in their variable regions. With this first glimpse of an Ig repertoire at the LC and HC levels, the presence of distinct proteoforms shows that single clone resolution is possible, with some present at high titer (i.e., ∼500 ng/100 uL plasma). The current dynamic range for detecting different clones that evolve after VDJ recombination is approximately 100 (**Supplementary Fig. 5**).

Given that Ig-MS produced a new data type, we created two new metrics. The Ion Titer (IT) is similar to an ELISA titer and uses the intensity of the LC of the standard mAb CR3022 as a reference. It combines intensity of all other LC peaks relative to the mAb CR3022 and ranges from 0 to 100 (**Equation 1**). The second is the Degree of Clonality (DoC), a measure of the complexity of all LC clones (proteoforms) in the mixture and ranges from 1 to infinity, with higher numbers reflecting the presence of a more significant number of antibodies in a more complex Ig-MS spectrum. For subject CS1, the calculated IT was 1.30, and the DoC was 3.64. We next sought to compare these Ig patterns and new metrics across patients and perform initial correlations with other COVID-19 antibody tests.

### Ig-MS of an initial cohort

We deployed Ig-MS workflows 1 and 2 to survey the Ig population reactive to RBD in a cohort of seven hospitalized patients with severe COVID-19, three outpatients with mild COVID-19 disease, and three healthy people who never had COVID-19 (**Supplementary Table 1**). The standard mAb CR3022 and commercial pooled plasma acquired before November 2019 served as positive and negative controls, respectively.

From workflow 1, the HCs and LCs are all shown in **Supplementary Fig. 6** for this cohort, while **Supplementary Fig. 7 and 8** present expansion of LC and HC regions, respectively. **Fig. 2** highlights the LC region for examples of three general groups of immunological responses: (**1**) high IT (30.11) and low DoC (2.36) as observed in one of the hospitalized patients (COVID-19_1) (**Fig. 2a**); (**2**) moderately high IT (14.87) and high DoC (7.25) as observed for the hospitalized patient COVID-19_3 (**Fig. 2b**); and (**3**) low IT (0.65) and low DoC (3.08) as observed for the outpatient COVID-19_6 (**Fig. 2c**).

**Fig. 2.**
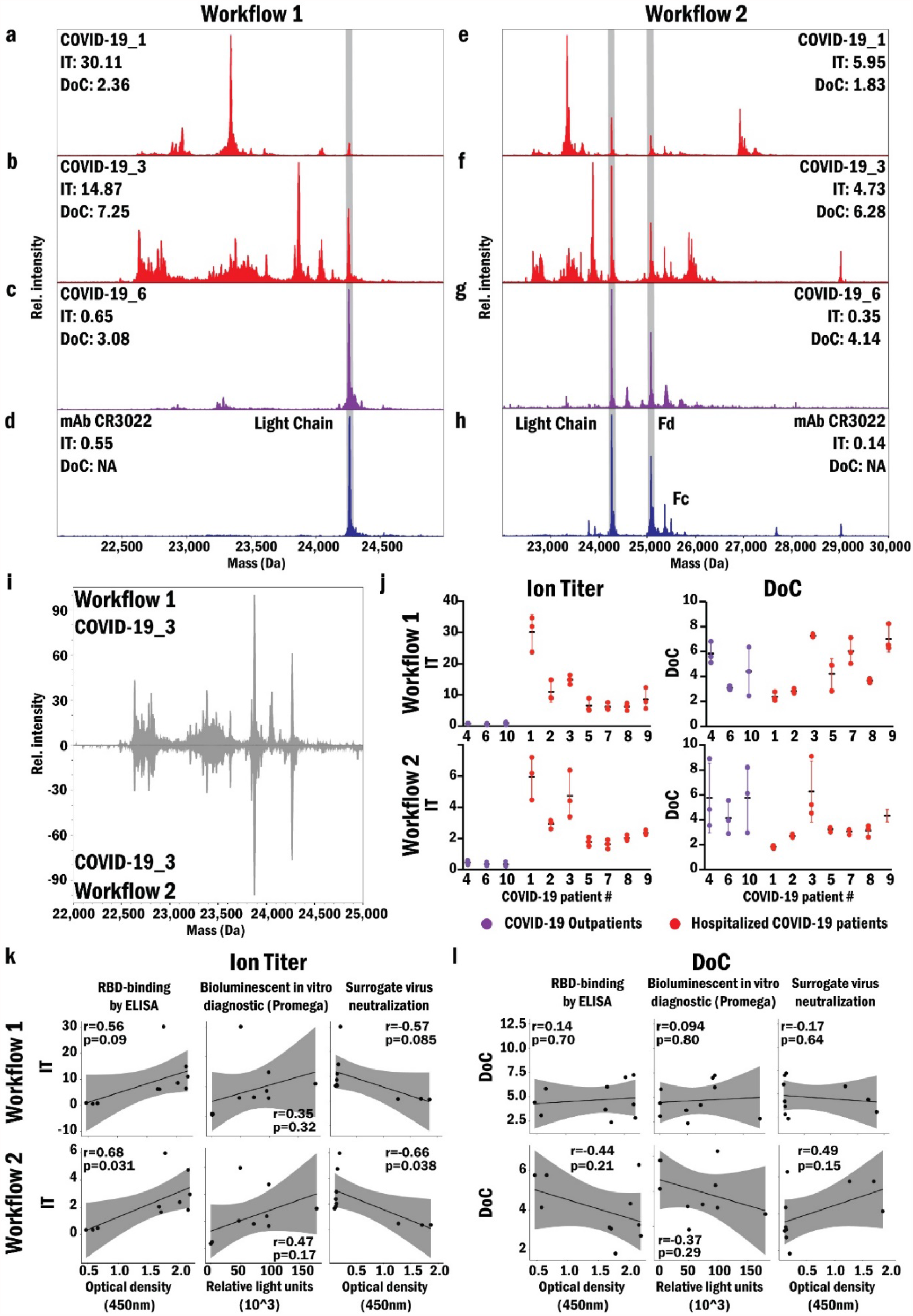
Ig-MS readouts from workflow 1 and 2 on a COVID-19 cohort. Results for the LC spectral region from **workflow 1** (a-d) and for the LC and Fd fragments from **workflow 2** (e-h); ion titers (IT), and degree of clonality (DoC) are shown for two hospitalized patients (a/e and b/f, red) and one outpatient (c/g, purple). The standard mAb (CR3022) that binds SARS-CoV-2-RBD (d/h, blue) was used as positive control (highlighted with gray vertical bar). i, Comparison of Ig-MS patterns between the LC of a single patient obtained with the two workflows. j, IT and DoC values obtained by both workflows of Ig-MS for three outpatients and seven hospitalized patients. Analyses were done in triplicate. Correlation of IT (k) and DoC (l) values from Ig-MS workflows with RBD-binding by ELISA, bioluminescent in vitro diagnostic (Promega), and surrogate virus neutralization. Shown are the Pearson correlation coefficient (r) and P-value (p).

The same 13 samples were processed using workflow 2, which generated the LC and Fd for readout by Ig-MS. **Supplementary Fig. 9** exhibits the obtained spectra for all samples. The spectra from patients COVID-19_1 (**Fig. 2e**), COVID-19_3 (**Fig. 2f**), and COVID-19_6 (**Fig. 2g**) displayed the same immunological response pattern as those obtained with workflow 1, showing the self-consistency of the two workflows.

Distinct proteoform masses representing all the LCs and Fds were clearly discernable in both workflows (**Fig. 2)**. The LC proteoforms have a mass range between 22-25 kDa (**Fig. 2**, to the left side of the standard LC highlighted in gray) and comprise the entire length of the LC, including the three CDRs (VL) and conserved region (CL). We can identify κ and λ LCs in the Ig-MS spectra based on the amino acid differences in the conserved regions. In **Fig. 2b**, the masses between 22.5 to 23 kDa belong to lambda LCs and those from 23 to 24 kDa to kappa LCs^18^. A correlation coefficient (**Equation 3**) was calculated and showed an average value of .95 for technical replicates. The same basic patterns of LC proteoforms were observed with both workflows applied to the same sample. For example, a comparison of LC proteoform patterns for patient COVID-19_3 (**Fig. 2i**) showed a correlation coefficient of 0.77. **Fig. 2j** displays the IT and DoC values for the subject’s samples. The IT values are significantly higher in hospitalized patients for both workflows (workflow1: F=12.64, 2 and 33 DF, p<0.001 and workflow 2: F=18.26, 2 and 33 DF, p<0.001) compared to outpatients, a result corroborated with standard serological tests from previous studies^19, 20^. No statistically significant differences were observed for the DoC metric in these groups.

Ion Titers were also calculated exclusively for LC regions and compared between workflow 1 and 2. Although differences occur in the range of ion titers observed between the two workflows, there was a strong correlation (R^2^ = 0.88) between them, indicating a systematic bias (**Supplementary Fig. 10**). We believe that this bias arises from the differences in sample and standard preparation. However, the average cross-correlation of the LC from the two studies is 0.78 ±0.04, indicating that the ratios among peaks and the relative proteoform amounts are conserved in both workflows of Ig-MS.

With workflow 2, we can analyze Fd proteoforms that include three CDRs from the HC (VH) and the constant region CH1 besides all possible individual variation present in the constant region (**Fig. 2**, to the right side of LC the standard Fd is highlighted in gray). This workflow captures the post-VDJ sequence recombination and dissects the Fc glycosylation pattern away from the intact HC observed in workflow 1, reducing molecular complexity. A general finding in this initial study is a greater degree of heterogeneity in the Fd relative to the LC from the same individual.

We next probed the correlation of ITs obtained with the two Ig-MS workflows with titers of anti-RBD antibodies quantified with ELISA^21^ and a commercial in vitro diagnostic using bioluminescence. We also determined the correlation between IT and neutralization efficiency obtained with a Spike-specific pseudovirus neutralization test (**Fig. 2k-l, Supplementary Fig. 11**, and **Supplementary Table 1**). Pearson’s correlation (r) (**Fig. 2k**) indicated a significant positive correlation of IT obtained with workflow 2 and the ELISA titers (r=0.68) and a negative correlation with surrogate neutralization (r=-0.66). The same tendency was observed for ITs determined with workflow 1 (r>0.55) but with *p*-values higher than 0.05. The correlation analysis of the ITs of both workflows with the bioluminescence was low (r≤0.47) and not significant. Furthermore, DoCs from both workflows did not correlate with the other assays (**Fig. 2l**), indicating that the number of different Ig signals produced by Ig-MS did not show a correlation with neutralization potential or overall titer in this initial study. For both workflows, there was a significant negative correlation between ITs and days after infection (**Supplementary Fig.12**). These results suggest that there is a reduction in the total amount of antibodies over time, but the general degree of complexity in the immune response does not change. Additionally, virus neutralization appears more related to the total amount of antibodies in the plasma than to clonal heterogeneity in the Ig repertoire.

### IgG glycosylation analysis

N-glycosylation at position 297 of the HC mediates the antibody’s effector functions. Further, the glycan moieties are highly variable and functionally relevant. To explore the IgG glycosylation pattern of our three study groups, we used bottom-up proteomics to quantify 17 different glycan structures (attached to the IgG peptide) and the unmodified peptide^22^. A Benjamini and Hochberg corrected ANOVA analysis on the results from the three groups indicated that seven fucosylated glycans (G2F, G0FN, G1FN, G2FN, G1FS1, G2FS1, and G2FS2) were differentially regulated. A second one-way ANOVA with Tukey’s multiple comparison test revealed that those fucosylated glycans were down-regulated in the hospitalized group compared to the uninfected group (**Supplementary Fig.13**).

Additionally, the average percent of the total fucosylated glycans was 53.9% ±17.7% for the hospitalized group and 95.5% ±0.8% for the uninfected individuals (**Supplementary Fig.14**). Similar results were previously reported^23^, and afucosylated IgG induces increased antibody-dependent cellular cytotoxicity by rising IgG-Fc receptor IIIa (FcγRIIIa) affinity^24^. Next, we tried to correlate these results to the Ig-MS readouts from workflow 1. Unfortunately, the high mass complexity observed in the HC region of workflow 1 generated from the different amino acid sequences and the multiple glycans structures pushes such analyses outside the scope of this initial study.

## Discussion

Antibody titers can be used to indicate the extent of immunity and disease severity for COVID-19^19, 20^. Ig-MS initial results showed that IT from the two workflows were self-consistent and correlated with traditional colorimetric/fluorimetric tests and a surrogate neutralization assay. The DoC metric and patterns of responses did not correlate with these assays, yet their variance in the human population needs to be determined. We observed significant differences in hospitalized patients versus outpatients as well as reduced IT overtime of convalescence, similar to previous reports^25^. With this initial report, Ig-MS proves functional, prompting three general use cases: 1) providing a new longitudinally-stable, multi-parametric correlate of protection, 2) indicating the course or stage of COVID-19 disease as a diagnostic or prognostic indicator, and 3) striating lots of convalescent plasma to quantify protective potential and better control plasma collection campaigns in this or future pandemics^26^. Additionally, antibody amounts and clonal variation can play a complementary role in vaccine campaigns that can strongly correlate to protective immunity (*i*.*e*., provide a reliable surrogate to neutralizing titers).

### Correlation with B cell Sequencing

Reports in the literature have begun to quantify the extent of convergence in the sequences of antibodies by deep sequencing of B cell receptors^27, 28^. These reports quantified responses across over 100 people and thousands of B cells, demonstrating the consistency of the stereotypical immune response, and are a great resource to identify potential therapeutic and prophylactic antibodies. Such convergence can also be detected by Ig-MS because it provides intact mass patterns that reflect the combination of LC and HC CDRs. This is unlike approaches using tryptic digestion of antibodies^8-11, 29^. Indeed, no technology can sequence whole, endogenous antibodies directly in an Ig repertoire, but >80% sequence coverage by direct fragmentation of LC and HC from monoclonal antibodies is possible today^30^.

### Future of COVID-19 Testing with Ig-MS

Moving forward, we will expand the cohort size into the hundreds to better define correlations with patient clinical metadata and other assays. Importantly, Ig-MS is adaptable just by switching the affinity resin, so use on RBD variants of Spike protein in virus strains (like the B.1.1.3 and P.1) will probe vaccination effectiveness and Ig-MS utility as a new correlate of protection in serology. In addition, there are reports of neutralizing epitopes in the *N*-terminal Domain (NTD) of Spike, and we can further probe this elucidated antibody repertoires^31^. Analysis of longitudinal samples from individuals infected with SARS-CoV-2 and after vaccination will track the immune response at high resolution^25, 32^. Ig-MS should also be applicable to IgA’s, IgMs, and other isotypes of immunoglobulins in the adaptive immune response.

In summary, we report a new and unique data type for human serology, using COVID-19 cases as the first example. Until now, no serological test was capable of accessing the relative abundance of each antibody generated against a specific antigen. Ig-MS is the first method capable of accessing amounts and relative abundance of antibodies simultaneously using a fundamental advance in MS of individual protein ions^16^ to create a unique display of the Ig repertoire of a human being at molecular resolution. Ig-MS successfully captured the clone populations of RBD-reactive immunoglobulins and showed promising data on a limited cohort of COVID-19 patients. In the future, a more automated form of Ig-MS will address larger cohorts, use 10-100 fold less sample, and extensively sequence CDR variable regions for comparison with methods for single B cell sequencing like Ig-seq^33^.

## Data Availability

Processed datasets utilized for the IgMS analyses can be found on the MassiVE repository, MSV000087529, after puplication. Custom compiled code used to process and create I2MS files is already available16. Additional desired software and data that support the findings of this study are available from the corresponding authors upon request.

## Acknowledgments

This study was funded by: the National Institute of Health under a grant from the National Institute of General Medical Sciences P41 GM108569 (N.L.K.); the Research Corporation (Grant no. 27372, N.L.K. and E.F.); Administrative Supplement to SBIR grant number R44 GM121130 (P.D.C.); Walder Foundation grant number SCI16 (J.R.S.); the NIH Office of Director award S10 OD025194 (P.D.C.); the Northwestern Medicine Dr. Michael M. Abecassis Transplant Innovation Endowment Grant; NCI CCSG P30 CA060553 (awarded to the Robert H. Lurie Comprehensive Cancer Center). The content of this paper is solely the responsibility of the authors and does not necessarily represent the official views of the National Institutes of Health.

## Author Contributions

Plasma isolation and storage by B.E.S., L.A.H, J.R.S. P.P.B, and M.K.A. Experiments developed and/or performed by R.D.M., B.J.S., J.O.K., E.F., F.N., V.B., J.P.M., B.D., C.L.J, H.S.S., P.D.C., B.K.C., B.B.M., Y.A.G., P.M.T., J.R.S., P.P.B, M.K.A, and N.L.K. Data processing and analysis by R.D.M, B.J.D., J.O.K, E.F., M.H., F.N., J.P.M., B.D., R.D.L., B.E., R.T.F., B.K.C., Y.A.G., P.M.T., and J.R.S. R.D.M., E.F., J.M.C., P.D.C., P.M.T., and N.L.K. collected funding support. R.D.M., B.J.D., E.F., and N.L.K. wrote and edited the manuscript. N.L.K. conceived the project, all authors critically reviewed and contributed to the construction of this project and manuscript in some fashion throughout the COVID-19 pandemic.

## Methods

### Patient Cohorts and Plasma Sampling

Throughout this work, plasma from convalescent COVID-19 donors and those in control groups were collected under IRB number 20032502-IRB01 by the clinical team at Rush University Medical Center. Patients were sampled post-infection 10 or more days after symptom onset. Plasma was isolated from the blood through centrifugation at 1,500 × *g* for 10 min. Plasma from a convalescent patient featuring a high titer of anti-SARS-CoV-2-RBD antibodies was purchased from AllCells (commercial sample 1 – CS1) and used as a standard positive control throughout the study. As a negative control/blank background, pooled plasma collected before the emergence of the SARS-CoV-2 pandemic was used (Fisher Scientific BP2657100 UNSPSC 12352207, purchased 05/05/2019).

### RBD-binding by ELISA

Hisorb Ni+ plates (Qiagen) were coated with 100 µL of His-tagged RBD (BEI) at a concentration of 2 µg/mL overnight at 4°C. Plates were blocked 100 µl per well of 3% non-fat milk prepared in PBS with 0.1% Tween 20 (PBST) was added to the plates at room temperature for 1 hr. Next, heat-inactivated plasma from COVID-19 patients was diluted 1:10 in PBS and added at 100 µL per well for 2 hr at RT. Plates were washed with PBS-T 3 times, followed by incubation with secondary anti-human IgG Fc HRP (1:4000) for 1 h. Plates were washed 3 times with PBS-T followed by the addition of TMB substrate for 10 min. The reaction was stopped using 3 M HCl and read at OD 450 nm on the Biotek Cytation 3.

### SARS-CoV-2 Surrogate Virus Neutralization Assay

COVID-19 patient plasma samples were heat-inactivated at 56°C for one hour. Following heat inactivation, samples were diluted at a volume ratio of 1:9 in sample dilution buffer and mixed with a 1:1000 HRP-conjugated RBD solution in HRP dilution buffer at a volume ratio of 1:1 and incubated at 37°C for 30 minutes. Following incubation, samples and kit-provided controls were added to an ACE2-coated 96-well plate, 100 µL/ well in duplicate, and incubated at 37°C for 15 minutes. The plate was then washed four times with 1X wash solution and incubated with 100 µL/well TMB solution in the dark at room temperature for 15 minutes, followed by the addition of 50 µL/well reaction stop solution. The absorbance was then read on a biotek Cytation 3 plate reader at 450 nm. The protocol is adapted from GenScript SARS-CoV-2 Surrogate Virus Neutralization Test Kit (Cat. No. L00847-A), and all reagents used were provided in the test kit.

### Recombinant Monoclonal Antibody

Human recombinant monoclonal Anti-SARS-CoV-2 antibody, mAb CR3022 produced in *Nicotiana bethamiana* (tobacco plant) was obtained from Novici Biotech LLC (Lot NCV_051520B) as a 1.0 mg/mL solution in PBS (pH 7.2). This reagent binds RBD of the Spike protein from SARS-CoV-2 and is also available through BEI Resources, catalog number NR-53876. The recombinant mAb was stored at 4°C prior to use.

### Recombinant expression of SARS-CoV-2 spike protein receptor-binding (RBD) domain

The plasmid pCAGGS SARS-CoV-2 RBD comprises an N-terminal signal sequence, amino acids 319-541 of the spike protein from SARS-CoV-2 (the receptor-binding domain, RBD), and a C-terminal 6-His tag^34^. This vector was obtained from BEI Resources (BEI NR-52309) and expressed recombinantly using the Expi293 Expression System as follows: Expi293F™ cell culture (1 L total) was maintained in a 37°C incubator with ≥80% relative humidity, 8% CO_2_ on an orbital shaker platform and sub-cultured at cell density 3-5 × 10^6^ viable cells/mL. One day before transfection the cells were seeded to a final density of 2.5 × 10^6^ viable cells/mL and allowed to grow overnight. On the day of transfection, the culture was diluted to 3 × 10^6^ viable cells/mL with fresh, prewarmed Expi293™ Expression Medium. The transfection was performed using ExpiFectamine™ 293 Transfection Kit. Briefly, DNA/Opti-MEM™ I and ExpiFectamine™ 293/Opti-MEM™ I mixtures were prepared separately and incubated at room temperature for 5 min. These mixtures were then combined, and the total complexation mixture was incubated at room temperature for another 20 min. after which it was slowly mixed into the cell culture to initiate transfection. 18 h post-transfection ExpiFectamine™ 293 Transfection Enhancer 1 and ExpiFectamine™ 293 Transfection Enhancer 2 were added to the culture. The cell culture supernatant was collected after 6 days and prepared for purification.

### Purification of SARS-CoV-2 RBD

Purification was performed on AKTAxpress (GE Healthcare Life Science) FPLC purification system. Clarified cell culture supernatant was loaded onto HisTrap FF 5 mL column (GE Healthcare Life Science, Cat #17-5255-01). The column was washed twice, once with binding buffer (10 mM Tris-HCl, 500 mM NaCl, pH 7.4) and then with binding buffer + 12.5 mM imidazole to remove unspecifically bound material. Finally, bound RBD protein was eluted off the column with elution buffer (10 mM Tris-HCl, 500 mM NaCl, 500 mM imidazole, pH 7.4). Column loading, washes, and elution were all performed at a 3 mL/min flow rate. After elution, the collected fraction was buffer exchanged into 1x PBS.

### RBD validation by bottom-up proteomics

Fifty μg of RBD was acetone/TCA precipitated with 8 volumes of cold acetone and one volume of trichloroacetic acid overnight at -20°C. After washing the pellet with ice-cold acetone, the resulting protein pellet was resuspended into 50 μL of 8 M urea in 400 mM ammonium bicarbonate, pH 7.8, reduced with 4 mM dithiothreitol at 50°C for 30 min., and cysteines were alkylated with 18 mM iodoacetamide in the dark for 30 min. The solution was then diluted to <2 M urea (final concentration), and trypsin (Promega) was added at a final trypsin/protein ratio of 1:50 prior to overnight incubation at 37°C with shaking. The resulting peptides were desalted using solid-phase extraction on a Pierce C18 Spin column and eluted in 80 μL of 80% acetonitrile (ACN) in 0.1% formic acid (FA). After lyophilization, peptides were reconstituted with 5% ACN in 0.1% FA.

Peptides were analyzed by LC-MS/MS using a Dionex UltiMate 3000 Rapid Separation nanoLC and a Q Exactive™ HF Hybrid Quadrupole-Orbitrap™ Mass Spectrometer (Thermo Fisher Scientific Inc, San Jose, CA). Approximately 1 μg of peptide sample was loaded onto the trap column, which was 150 μm x 3 cm in-house packed with 3 μm ReproSil-Pur® C18 beads (Maisch, GmbH, Germany). The analytical column was a 75 μm x 10.5 cm PicoChip column packed with 3 μm ReproSil-Pur® C18 beads (New Objective, Inc. Woburn, MA). The flow rate was kept at 300 nL/min. Solvent A was 0.1% FA in water, and Solvent B was 0.1% FA in ACN. The peptides were separated on a 120 min. analytical gradient from 5% ACN/0.1% FA to 40% ACN/0.1% FA. The mass spectrometer was operated in a data-dependent mode. The source voltage was 2.40 kV, and the capillary temperature was 320°C. MS^1^ scans were acquired from 300-2000 *m/z* at 60,000 resolving power and automatic gain control (AGC) set to 3×10^6^ charges. The top 20 most abundant precursor ions in each MS^1^ scan were selected for fragmentation. Precursors were selected with an isolation width of 2 *m/z* and fragmented by Higher-energy collisional dissociation (HCD) at 30% normalized collision energy in the HCD cell. Previously selected ions were dynamically excluded from re-selection for 20 seconds. The MS^2^ minimum AGC was set to 1×10^3^.

Proteins were identified from the tandem mass spectra extracted by Xcalibur version 4.0. MS^2^ spectra were searched against the Spike protein RBD and SwissProt *Homo sapiens* database using Mascot search engine (Matrix Science, London, UK; version 2.7.0). All searches included carbamidomethyl cysteine as a fixed modification, oxidized methionine, deamidated asparagine/glutamine, and acetylated *N*-terminal as variable modifications. Three missed tryptic cleavages were allowed. The MS^1^ precursor mass tolerance was set to 10 ppm, and the MS^2^ tolerance was set to 0.05 Da. The search result was visualized by Scaffold v 5.0 (Proteome Software, INC., Portland, OR). A 1% false discovery rate cutoff was applied at the peptide level. Only proteins with a minimum of two unique peptides above the cutoff were considered identifications.

### Fabrication of RBD-loaded magnetic beads

Recombinantly-expressed RBD was covalently loaded onto Dynabeads® MyOne™ Carboxylic Acid beads (Thermo 65011, Thermo 65012) according to the manufacturer-provided protocol for “Two-Step Coating Procedure using NHS”. In brief, for a single preparation, 1.5 mL of bead suspension was dispensed and washed twice with 1.5 mL of 25 mM MES, pH 6.0 for 10 min. at room temperature. Following washes, bead chemistry was activated by suspending the beads into 1.5 mL of freshly-prepared 50 mg/mL N-Hydroxysuccinimide (NHS), mixing in 1.5 mL of freshly-prepared 50 mg/mL 1-Ethyl-3- (3dimethylaminopropyl) carbodiimide (EDC), and incubating for 30 min. at room temperature. Following activation, the beads were again washed twice with 1.5 mL of 25 mM MES, pH 6.0 for 10 min. at room temperature. Activated beads were suspended into 1 mL 25 mM MES, pH 6.0 to which 1.5 mg of RBD protein was added. The sample was mixed and incubated for 30 min. at room temperature. Next, the beads were pulled down, the supernatant was discarded, and loading was quenched by incubating the beads in 1 mL of 50 mM Tris, pH 6.8, for 15 min. at room temperature. After quenching, beads were washed four times in 1 mL 1x PBS + 0.1% human serum albumin (HSA), and finally suspended into 3 mL 1x PBS + 0.1% HSA for storage at 4°C.

### Enrichment of RBD-reactive antibodies from patient samples

Antibody pull-downs were assembled by combining 100 µL of patient plasma/serum with 35 µL of RBD-loaded bead suspension and diluting to a volume of 1 mL with 1x TBS. Assembled pull-downs were incubated overnight at 4°C with end-over-end mixing. After this, incubation beads were pulled down, supernatant was removed, and beads were resuspended into 1 mL wash buffer (1x TBS + 0.1% TWEEN + 1% NP-40 + 1% NP-40 substitute). Suspensions were transferred onto a KingFisher Flex for additional 4 washes in 1 mL wash buffer, 2 washes in 1 mL 1x TBS, and a 30 min. incubation in 100 µL 100 mM glycine, pH 11.5 + 0.1% sodium deoxycholate at 37°C to elute antibodies associated with bead-bound RBD. For workflow 2, Ig-pull-downs included an on-bead IdeS digestion of RBD-binding antibodies. This involved an additional incubation step between the first and second 1x TBS washes, during which beads were incubated in 100 µL of 50 mM sodium phosphate + 150 mM sodium chloride (pH 6.6) in the presence of 200 U IdeS enzyme (Promega V751A) for 3.5 h at 37°C.

### Preparation of enriched antibodies for individual ion mass spectrometry

Following elution, 100 ng of mAb CR3022 standard antibody was added to each elution fraction to serve as an internal standard across all samples. For samples including an IdeS digest step, the mAb CR3022 standard underwent an in-solution IdeS digestion according to manufacturer protocols prior to being added to pull-down elution fractions. After supplementation with standard antibody, each fraction was combined with 160 µL 8M urea and 25 µL 1M TCEP, mixed, and set to incubate for 1 h at room temperature to facilitate the complete denaturation of the Ig-RBD and permit the complete reduction of all inter- and intrachain disulfide bonds. Following incubation, reduced antibody fragments were cleaned via methanol-chloroform-water precipitation as has been previously described^35, 36^.

### Visualization of RBD-binding antibodies via western blot

Fractions of interest were combined with Bolt™ loading buffer (final concentration 1x, Thermo Fisher B0007) and dithiothreitol (DTT, final concentration 100 mM) and incubated at 100°C for 10 min. Boiled samples were loaded onto a 4-12% Bolt™ Bis-Tris Plus polyacrylamide gel, which was run in a Mini Gel Tank (Thermo Fisher A25977) using MES running buffer for 1 h at 120 V. Next, the gel was trimmed, and proteins were transferred to a nitrocellulose membrane using iBlot™ 2 nitrocellulose transfer stacks (Thermo Fisher IB23001) on an iBlot™ 2 Dry Blotting System following manufacturer instructions. The transfer method used was the templated method P3 (20 V for 7 min.). After transfer, the membrane was blocked using 1x TBS + 0.05% TWEEN + 5% milk for 1 h at room temperature with mixing. After 1 h, Goat anti-Human IgG (H+L) HRP conjugate (Thermo Fisher 31410) was added directly to the milk at a 1:20000 dilution and the membrane was moved to 4°C to incubate overnight. The next day, the milk + antibody mixture was discarded, and the membrane was washed 3x in 1x TBS + 0.05% TWEEN for 5 min. at room temperature with mixing. Finally, the blot was imaged on an iBright CL1000 imager (Thermo Fisher) using 800 µL of Immobilon Classico Western HRP substrate (Millipore Sigma WBLUC0500).

### Western blot identification of enriched antibody isotypes

An enrichment was performed as described in the section “*Enrichment of RBD-reactive antibodies from patient samples”* above to screen for the presence of individual human antibody isotypes (IgG1, IgG2, IgG3, IgG4, IgA, IgD, IgE, and IgM). In this case, after elution, the fraction was divided into 8 equal parts. Positive controls were assembled for each isotype being examined, with each control consisting of 250 ng of a commercially-obtained purified antibody of that isotype (IgG1: Sigma AG502; IgG2: Sigma I5404; IgG4: Sigma I4640; IgA: Sigma I4036; IgD: Sigma 401164; IgE: Sigma 401152; IgM: Sigma I8260). Negative controls for each isotype were assembled by combining 300 ng of all commercially obtained purified antibody isotypes omitting the specific isotype that the control was. Eluted material, positive, and negative controls were run in polyacrylamide gels, and proteins were transferred to nitrocellulose and blocked as described above. After blocking, each membrane was introduced to a primary antibody that specifically recognized the isotype of the antibodies being examined on that membrane (IgG1: AbCam ab108969; IgG2: AbCam ab134050; IgG3: AbCam ab109761; IgG4: AbCam ab109493; IgA: AbCam ab124716; IgD: AbCam ab124795; IgE: AbCam ab195580; IgM: AbCam ab134159) mixed with 1x TBS + 0.05% TWEEN + 5% milk. Primary antibodies were present at a dilution of 1:10000. Membranes were incubated in primary antibodies overnight at 4°C with mixing. The next day, primary antibody + milk was discarded, and all membranes were washed 3x in 1x TBS + 0.05% TWEEN for 5 min. at room temperature with mixing. After washing, all membranes were submerged into 1x TBS + 0.05% TWEEN + 5% milk to which secondary antibody (Gt-anti-Rb-HRP (Sigma AP307P) had been added at a dilution of 1:5000. Membranes incubated in secondary antibody for 1 h at room temperature with mixing, after which they were washed 3x in 1x TBS + 0.05% TWEEN for 5 min. at room temperature with mixing and imaged as described above.

### Anti-RBD antibody quantification

Titers of antibodies from patient plasma responsive to SARS-CoV-2 RBD were measured using Lumit™ Dx SARS-CoV-2 Immunoassay (Promega VB1080). Plasma from each patient was diluted 1:10 in 1x TBS and incubated at 56°C for 1 hr to inactivate any potential pathogens remaining. Following heat inactivation, samples were used as input and analyzed in triplicate using the assay following manufacturer protocols.

### Individual ion mass spectrometry

After precipitation, reduced antibody samples were redissolved in 200 μL of a 40% ACN and 0.5% acetic acid. A total of 70 μL of the sample was sprayed through an Ion Max Source (Thermo Fisher Scientific) fitted with a HESI II probe at a flow rate of 1 μL/min delivered by a PAL3 robot (CTC Analytics) and analyzed by a Q Exactive Plus mass spectrometer (Thermo Fisher Scientific)^37, 38^. Instrumental conditions included eFT off, pressured setting of 0.5, 1kV orbitrap central electrode voltage, a spray voltage of 3.0 to 4 kV, sheath gas of 2 L/min., an in-source collision-induced-dissociation value of ∼15 V, and a source temperature of 320°C. Injection times ranged from 1-400 ms and were optimized on a per-sample basis to collect hundreds to thousands of individual ions (depending on spectral complexity) per acquisition event to perform I^2^MS analysis. Data were acquired from 650-2,500 *m/z* range for 70 min. using 140,000 of resolving power (at 200 *m/z*).

Previously, I^2^MS analysis has been demonstrated using an Orbitrap mass analyzer^16, 39, 40^. Briefly, I^2^MS is a novel approach capable of discerning the mass profile of highly complex mixtures not amenable to venerable approaches used in protein mass spectrometry. Rather than measuring in the mass-to-charge (*m/z*) domain, I^2^MS accurately determines the charge on each individual ion collected through a process called STORI plot analysis. STORI plot analysis, the determination of the slope of individual ion signal accumulation, was completed on each ion as a function of its specific frequency^16^. To simplify this process and reduce file sizes, only time-domain signals at specific frequencies where individual ions occurred, called STORI files, were recorded on an acquisition-to-acquisition basis. Each acquired STORI file was then processed to accurately determine the *m/z* and charge for every individual ion signal detected.

In order to evaluate the lower limits of the GIDI-UP platform, we analyzed NIST antibody standard in a serial dilution series. All samples were infused at 1 µL/min. for 50 min. such that the same number of transients would be acquired for each experiment. Additionally, the injection time was scaled inversely to the sample concentration in order to maintain a constant per-cycle injection of 12.5 pg. For example, the 500 nM run used an injection time of 10 ms, and the 100 nM run used an injection time of 50 ms.

### Proteoform Quantification for Light Chain, Fd, and Heavy Chain

I^2^MS STORI files were processed to create mass spectra as .*mzml* files. Briefly, STORI files containing single ion peak information and transient sections are processed using a Short-time Fourier transform (STFT) to assign slopes to single ions^41^. After slope assignment, charges were assigned to individual ion using an iterative voting algorithm, and spectra were generated using a Normal kernel density estimate (KDE) and exported as either profile or centroided .*mzml* files.

### Calculation of Ig-MS Ion Titer, Degree of Clonality and Spectral Correlation Coefficients

Ig-MS Ion Titers were calculated using a custom script, in which .*mzml* I^2^MS files were centroided and spectra divided into regions; the Light Chain (LC) region, the regions of the Standard peaks, and either the Fd fragment or Heavy Chain (HC) regions for samples that were reduced with and without IdeS digestion, respectively. An average noise was calculated over these regions, and the sum was subtracted from the intensity for the region’s total^42^. The titers were obtained by determining the ratio between the sum of the standard peak regions and the regions for the LC and Fd/HC regions, divided by the value of the spike-in standard, as shown in **Equation 1**.

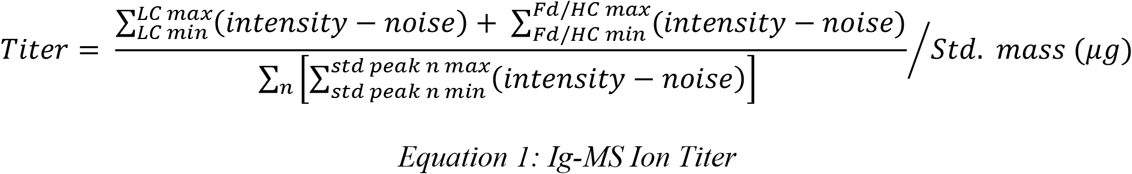

The degree of clonality (DoC) was calculated by centroiding profile spectra for a given mass window corresponding to the LC region. Firstly, the highest centroid peak is determined and using an averagine distribution, a window is created around the peak^43^. To account for simple adducts (+Na and -H_2_O) the window was extended +39 and – 18 Da. The intensities in this range are summed and divided by the total sum of all intensities in the LC region and inverted yield the DoC, as in **Equation 2**. This metric takes a value of one to infinity, with the higher the number, the more complex the spectrum.

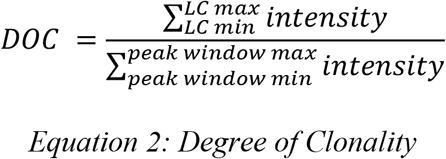

Spectral correlation coefficients were calculated for two spectra by taking centroided spectra and creating a padded array of equal length for each. Each peak in each centroided spectra was fitted to a Normal KDE and summed into the padded arrays, yielding two gaussian fitted spectra of equal length with indexes corresponding to the same mass. Calculating the Cosine Similarity using **Equation 3**, yielded the spectral correlation coefficients of the two spectra.

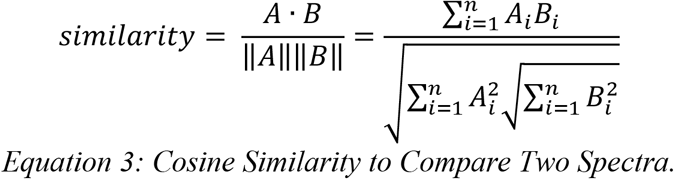

### IgG Antibody glycan analysis

Reduced IgG purified from plasma were separated on an SDS-PAGE gel and the band correspondent to the heavy chain was cut and subjected to in-gel digestion as follows: gel bands were washed in 100 mM Ammonium Bicarbonate (AmBic)/Acetonitrile (ACN) and reduced with 10 mM dithiothreitol at room temperature for 45 min. Cysteines were alkylated with 50 mM iodoacetamide in the dark for 45 min. at room temperature. Finally, gel bands were washed in 100 mM AmBic/ACN prior to adding 600 ng Lys-C for overnight incubation at room temperature. Following digest, supernatants containing peptides were transferred into new tubes. Gel pieces were washed at room temperature for 10 min. with gentle shaking, in 50% ACN/5% FA, and supernatants were combined with peptide solutions. This wash was repeated each by 80% ACN/5% FA, and 100% ACN, and all supernatants was saved. Pooled supernatants were then subject to speedvac drying. After lyophilization, peptides were reconstituted with 5% ACN/0.1% FA in water.

Peptides were analyzed by LC-MS/MS using a Dionex UltiMate 3000 Rapid Separation nanoLC and a Q Exactive™ HF Hybrid Quadrupole-Orbitrap™ Mass Spectrometer (Thermo Fisher Scientific Inc). The peptide samples were loaded onto the trap column, which was 150 μm x 3 cm in-house packed with 3 µm C18 beads. The analytical column was a 75 µm x 10.5 cm PicoChip column packed with 3 µm C18 beads (New Objective). The flow rate was kept at 300 nL/min. Solvent A was 0.1% FA in water and Solvent B was 0.1% FA in ACN. The peptide was separated on a 60 min. analytical gradient from 5% to 50% of Solvent B. The mass spectrometer was operated in the Full MS scan. The source voltage was 2.30 ∼ 2.50 kV and the capillary temperature 320°C. Full MS scans were acquired from 400-2000 *m/z* at 60,000 resolving power and automatic gain control (AGC) set to 3×10^6^.

MS data were processed using Skyline (Version 20.2). The integration and correction for the chromatographic peaks of 18 glycans were performed manually. Three of the most intense precursor ions of each glycan were selected, summed, and exported as the quantitative value of the corresponding glycan. Total ion intensities were used to generate the plots.

### Statistical analysis

ANOVA statistics were calculated using SAS (SAS Institute, Cary, NC). Person’s correlations were calculated using the function “cor.test” and method “pearson” on RStudio v3.1.1073. Violin and dot plots were generated with GraphPad Prism 9.0.2 and that same software was used for calculating the statistical significance by one-way ANOVA with Tukey’s multiple comparison test (* p<0.05; ** p<0.01).

### Data and software availability

Processed datasets utilized for the IgMS analyses can be found on the MassiVE repository, MSV000087529, after puplication. Custom compiled code used to process and create I^2^MS files is already available^16^. Additional desired software and data that support the findings of this study are available from the corresponding authors upon request.

### Methods acknowledgments

RBD was expressed by the Northwestern Recombinant Protein Production Core Facility. The reagent was produced under HHSN272201400008C and obtained through BEI Resources, NIAID, NIH: Vector pCAGGS Containing the SARS-Related Coronavirus 2, Wuhan-Hu-1 Spike Glycoprotein Receptor-Binding Domain (RBD), NR-52309. IgG glycosylation and bottom-up proteomics were measured by the Northwestern Proteomics Core Facility.

## References

1. Zhou, P. et al. A pneumonia outbreak associated with a new coronavirus of probable bat origin. Nature 579, 270–273 (2020).

2. Zhou, Y. et al. Comorbidities and the risk of severe or fatal outcomes associated with coronavirus disease 2019: A systematic review and meta-analysis. Int J Infect Dis 99, 47–56 (2020).

3. Ahmad, F.B., Cisewski, J.A., Minino, A. & Anderson, R.N. Provisional Mortality Data - United States, 2020. MMWR Morb Mortal Wkly Rep 70, 519–522 (2021).

4. Wang, P. et al. Antibody resistance of SARS-CoV-2 variants B.1.351 and B.1.1.7. Nature (2021).

5. Wu, J. et al. SARS-CoV-2 infection induces sustained humoral immune responses in convalescent patients following symptomatic COVID-19. Nat Commun 12, 1813 (2021).

6. Kreer, C. et al. Longitudinal Isolation of Potent Near-Germline SARS-CoV-2-Neutralizing Antibodies from COVID-19 Patients. Cell 182, 1663–1673 (2020).

7. Rogers, T.F. et al. Isolation of potent SARS-CoV-2 neutralizing antibodies and protection from disease in a small animal model. Science 369, 956–963 (2020).

8. Wine, Y. et al. Molecular deconvolution of the monoclonal antibodies that comprise the polyclonal serum response. Proc Natl Acad Sci U S A 110, 2993–2998 (2013).

9. Georgiou, G. et al. The promise and challenge of high-throughput sequencing of the antibody repertoire. Nat Biotechnol 32, 158–168 (2014).

10. Cheung, W.C. et al. A proteomics approach for the identification and cloning of monoclonal antibodies from serum. Nat Biotechnol 30, 447–452 (2012).

11. Tran, N.H. et al. Complete De Novo Assembly of Monoclonal Antibody Sequences. Sci Rep 6, 31730 (2016).

12. Wheatley, A.K. et al. Evolution of immune responses to SARS-CoV-2 in mild-moderate COVID-19. Nat Commun 12, 1162 (2021).

13. Lee, J. et al. Molecular-level analysis of the serum antibody repertoire in young adults before and after seasonal influenza vaccination. Nat Med 22, 1456–1464 (2016).

14. Smith, L.M., Kelleher, N.L. & Consortium for Top Down, P. Proteoform: a single term describing protein complexity. Nat Methods 10, 186–187 (2013).

15. He, L. et al. Analysis of Monoclonal Antibodies in Human Serum as a Model for Clinical Monoclonal Gammopathy by Use of 21 Tesla FT-ICR Top-Down and Middle-Down MS/MS. J Am Soc Mass Spectrom 28, 827–838 (2017).

16. Kafader, J.O. et al. Multiplexed mass spectrometry of individual ions improves measurement of proteoforms and their complexes. Nat Methods 17, 391–394 (2020).

17. Amanat, F. et al. A serological assay to detect SARS-CoV-2 seroconversion in humans. Nat Med 26, 1033–1036 (2020).

18. Barnidge, D.R. et al. Phenotyping Polyclonal Kappa and Lambda Light Chain Molecular Mass Distributions in Patient Serum Using Mass Spectrometry. Journal of Proteome Research 13, 5198–5205 (2014).

19. Klein, S.L. et al. Sex, age, and hospitalization drive antibody responses in a COVID-19 convalescent plasma donor population. J Clin Invest 130, 6141–6150 (2020).

20. Lynch, K.L. et al. Magnitude and Kinetics of Anti–Severe Acute Respiratory Syndrome Coronavirus 2 Antibody Responses and Their Relationship to Disease Severity. Clinical Infectious Diseases 72, 301–308 (2020).

21. Amanat, F. et al. A serological assay to detect SARS-CoV-2 seroconversion in humans. Nature Medicine 26, 1033–1036 (2020).

22. Gunn, B.M. et al. Enhanced binding of antibodies generated during chronic HIV infection to mucus component MUC16. Mucosal Immunology 9, 1549–1558 (2016).

23. Larsen, M.D. et al. Afucosylated IgG characterizes enveloped viral responses and correlates with COVID-19 severity. Science 371, eabc8378 (2021).

24. Ferrara, C. et al. Unique carbohydrate–carbohydrate interactions are required for high affinity binding between FcγRIII and antibodies lacking core fucose. Proceedings of the National Academy of Sciences (2011).

25. Lau, E.H.Y. et al. Neutralizing antibody titres in SARS-CoV-2 infections. Nat Commun 12, 63 (2021).

26. Klassen, S. et al. Convalescent Plasma Therapy for COVID-19: A Graphical Mosaic of the Worldwide Evidence. SSRN (2021).

27. Galson, J.D. et al. Deep Sequencing of B Cell Receptor Repertoires From COVID-19 Patients Reveals Strong Convergent Immune Signatures. Frontiers in Immunology 11 (2020).

28. Robbiani, D.F. et al. Convergent antibody responses to SARS-CoV-2 in convalescent individuals. Nature 584, 437–442 (2020).

29. Guthals, A. et al. De Novo MS/MS Sequencing of Native Human Antibodies. J Proteome Res 16, 45–54 (2017).

30. Fornelli, L. et al. Accurate Sequence Analysis of a Monoclonal Antibody by Top-Down and Middle-Down Orbitrap Mass Spectrometry Applying Multiple Ion Activation Techniques. Anal Chem 90, 8421–8429 (2018).

31. Liu, L. et al. Potent neutralizing antibodies against multiple epitopes on SARS-CoV-2 spike. Nature 584, 450–456 (2020).

32. Yamayoshi, S. et al. Antibody titers against SARS-CoV-2 decline, but do not disappear for several months. EClinicalMedicine 32, 100734 (2021).

33. Lopez-Santibanez-Jacome, L., Avendano-Vazquez, S.E. & Flores-Jasso, C.F. The Pipeline Repertoire for Ig-Seq Analysis. Front Immunol 10, 899 (2019).

34. Stadlbauer, D. et al. SARS-CoV-2 Seroconversion in Humans: A Detailed Protocol for a Serological Assay, Antigen Production, and Test Setup. Curr Protoc Microbiol 57, e100 (2020).

35. Toby, T.K. et al. A comprehensive pipeline for translational top-down proteomics from a single blood draw. Nat Protoc 14, 119–152 (2019).

36. Wessel, D. & Flugge, U.I. A method for the quantitative recovery of protein in dilute solution in the presence of detergents and lipids. Anal Biochem 138, 141–143 (1984).

37. Skinner, O.S. et al. An informatic framework for decoding protein complexes by top-down mass spectrometry. Nat Methods 13, 237–240 (2016).

38. Park, H.-M. et al. Novel Interface for High-Throughput Analysis of Biotherapeutics by Electrospray Mass Spectrometry. Analytical chemistry 92, 2186–2193 (2020).

39. Kafader, J.O. et al. Individual Ion Mass Spectrometry Enhances the Sensitivity and Sequence Coverage of Top-Down Mass Spectrometry. Journal of Proteome Research 19, 1346–1350 (2020).

40. McGee, J.P. et al. Isotopic Resolution of Protein Complexes up to 466 kDa Using Individual Ion Mass Spectrometry. Analytical Chemistry 93, 2723–2727 (2021).

41. Kafader, J.O. et al. STORI Plots Enable Accurate Tracking of Individual Ion Signals. J Am Soc Mass Spectrom 30, 2200–2203 (2019).

42. Horn, D.M., Zubarev, R.A. & McLafferty, F.W. Automated reduction and interpretation of high resolution electrospray mass spectra of large molecules. J Am Soc Mass Spectrom 11, 320–332 (2000).

43. Senko, M.W., Beu, S.C. & McLafferty, F.W. Determination of monoisotopic masses and ion populations for large biomolecules from resolved isotopic distributions. Journal of the American Society for Mass Spectrometry 6, 229–233 (1995).

