## Supplementary Information for "Next-generation Serology by Mass Spectrometry: Readout of the SARS-CoV-2 Antibody Repertoire"

**Supplementary Table 1. Correlation of Ig-MS metrics with patient metadata.** Average titers were determined with ELISA-based serological assays and surrogate virus neutralization titers from 10 COVID-19 convalescent patients. IgMS metrics for workflow 1 and workflow 2. Ion titers ranges from 0 to 100 and degree of clonality (DoC) from 1 to infinity. Both metrics are the average of 3 replicates.

| Metadata | | | Titers | | | Workflow 1 | | Workflow 2 | |
|---|---|---|---|---|---|---|---|---|---|
| Patient | Hospitalized | Days from test to blood draw | ELISA | Promega | Surrogate Neutralization | Ion Titer | DoC | Ion Titer | DoC |
| COVID-19_1 | Yes | 20 | 1.78 | 50345 | 0.20 | 30.11 | 2.36 | 5.95 | 1.83 |
| COVID-19_2 | Yes | 10 | 2.19 | 179569 | 0.14 | 10.99 | 2.82 | 2.94 | 2.71 |
| COVID-19_3 | Yes | 27 | 2.16 | 98789 | 0.16 | 14.87 | 7.25 | 4.73 | 6.28 |
| COVID-19_4 | No | 90 | 0.66 | 2471 | 1.27 | 0.83 | 5.84 | 0.46 | 5.76 |
| COVID-19_5 | Yes | 57 | 2.16 | 73400 | 0.13 | 6.51 | 4.22 | 1.80 | 3.26 |
| COVID-19_6 | No | 50 | 0.58 | 843 | 1.86 | 0.65 | 3.08 | 0.35 | 4.14 |
| COVID-19_7 | Yes | 67 | 1.71 | 98706 | 0.11 | 6.22 | 6.03 | 1.64 | 3.07 |
| COVID-19_8 | Yes | 71 | 1.68 | 47852 | 0.14 | 6.30 | 3.67 | 2.03 | 3.14 |
| COVID-19_9 | Yes | 60 | 2.02 | 95191 | 0.13 | 8.59 | 7.01 | 2.37 | 4.34 |
| COVID-19_10 | No | Not tested | 0.47 | 704 | 1.70 | 0.94 | 4.41 | 0.34 | 5.76 |
| Uninfected_1 | NA | NA | - | 635 | - | 0.22 | NA | 0.28 | NA |
| Uninfected_2 | NA | NA | - | 450 | - | 0.39 | NA | 0.37 | NA |
| Uninfected_3 | NA | NA | - | 517 | - | 0.38 | NA | 0.55 | NA |
| Standard mAb | NA | NA | - | - | - | 0.55 | NA | 0.14 | NA |

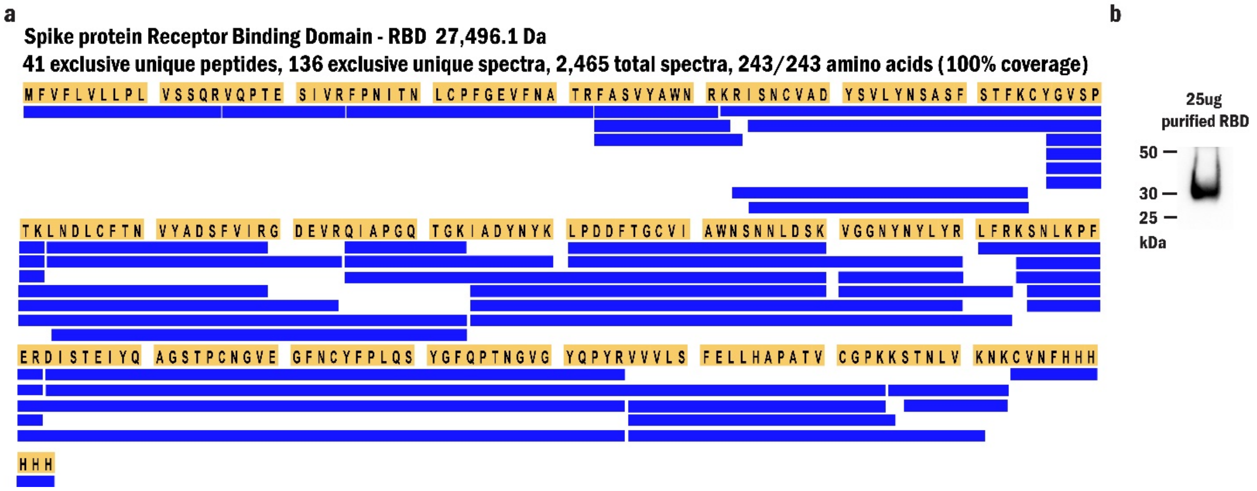


**Supplementary Fig. 1**. **Expression validation of RBD region from the SARS-CoV-2 Spike protein.** (a) A peptide map of Spike-RBD from bottom-up proteomics showing 100% sequence coverage. (b) Western-blot using the mAb CR3022 standard monoclonal antibody that recognizes RBD as primary antibody.

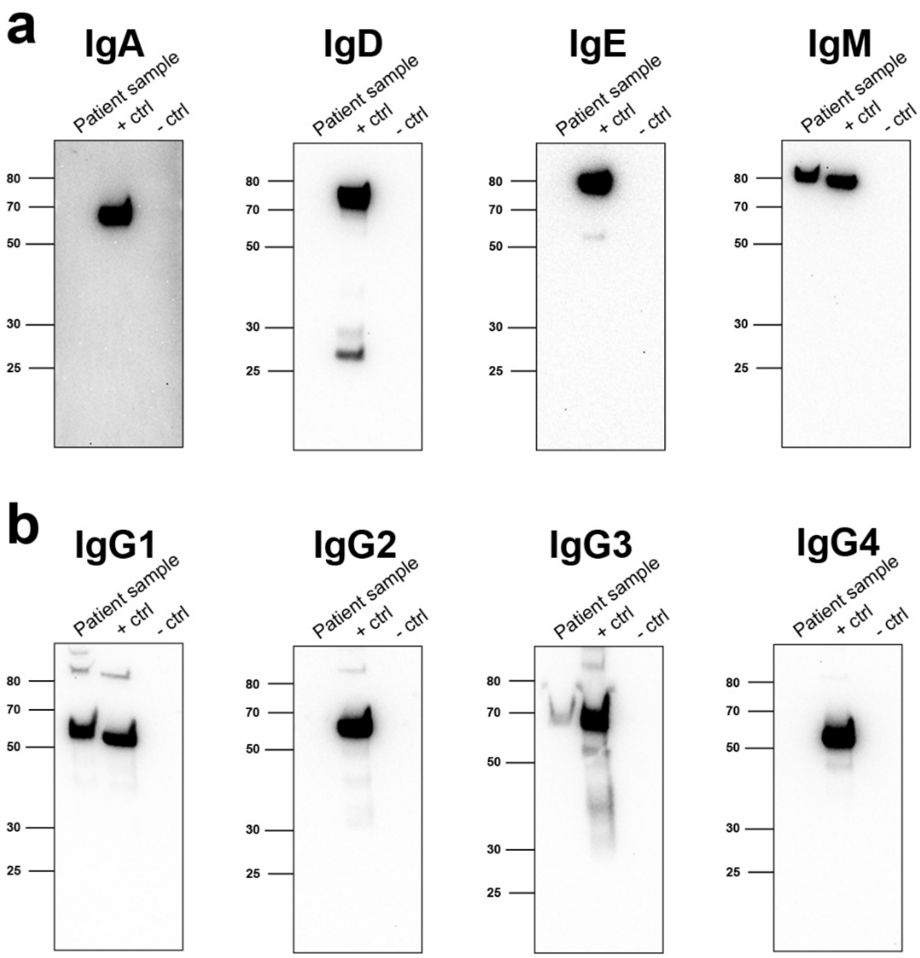


**Supplementary Fig. 2. Western blot showing isotypes and IgG subclasses of the antibodies enriched with Ig-MS assay.** RBD-specific antibodies were immunoprecipitated from the plasma of a COVID-19 convalescent patient with magnetic beads conjugated with the recombinant RBD protein and separated via SDS-PAGE. Proteins were transferred to nitrocellulose and probed with antibodies recognizing human (a) antibody isotypes IgA, IgD, IgE, and IgM or (b) IgG subclasses IgG1, IgG2, IgG3, and IgG4. Isotype-specific antibodies were used as positive controls, and pooled human isotype antibodies excluding the isotype of interest were used as negative controls.

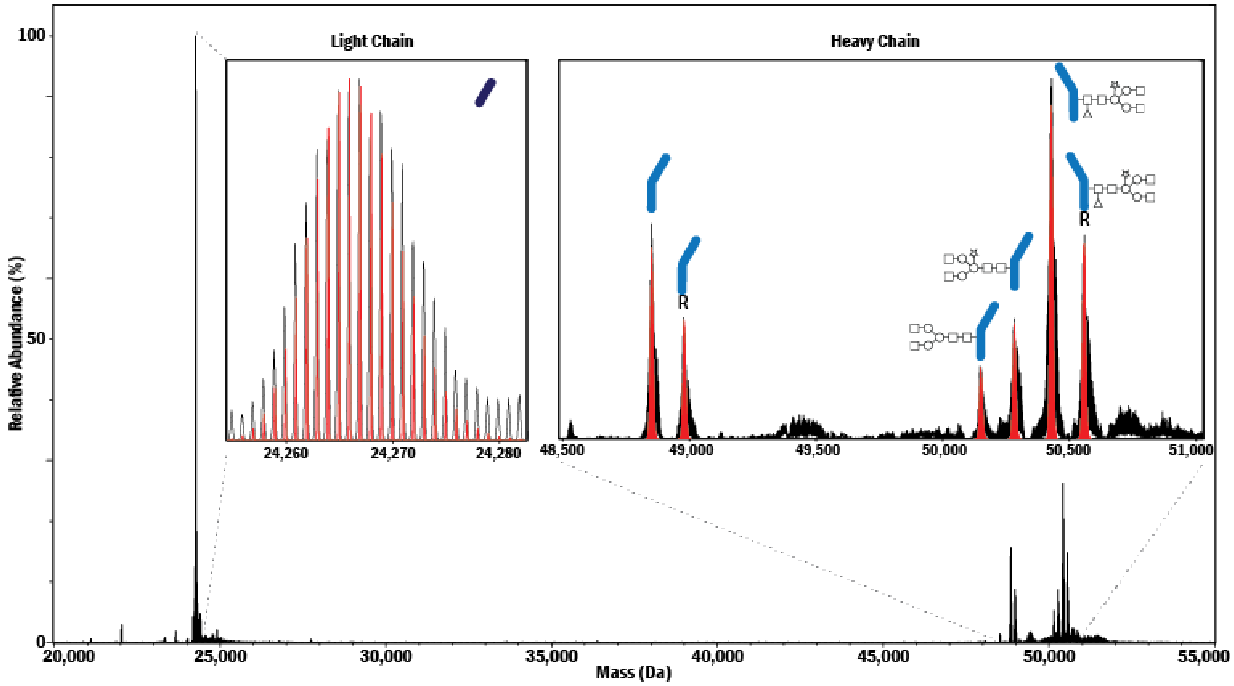


**Supplementary Fig. 3. Proteoform annotation of the CR3022 standard monoclonal antibody.** Mass spectrum acquired by Ig-MS in black and the theoretical mass distribution in red for the light chain and glycoproteoforms on the heavy chain of the antibody, produced in *Nicotiana bethamiana* (tobacco plant).

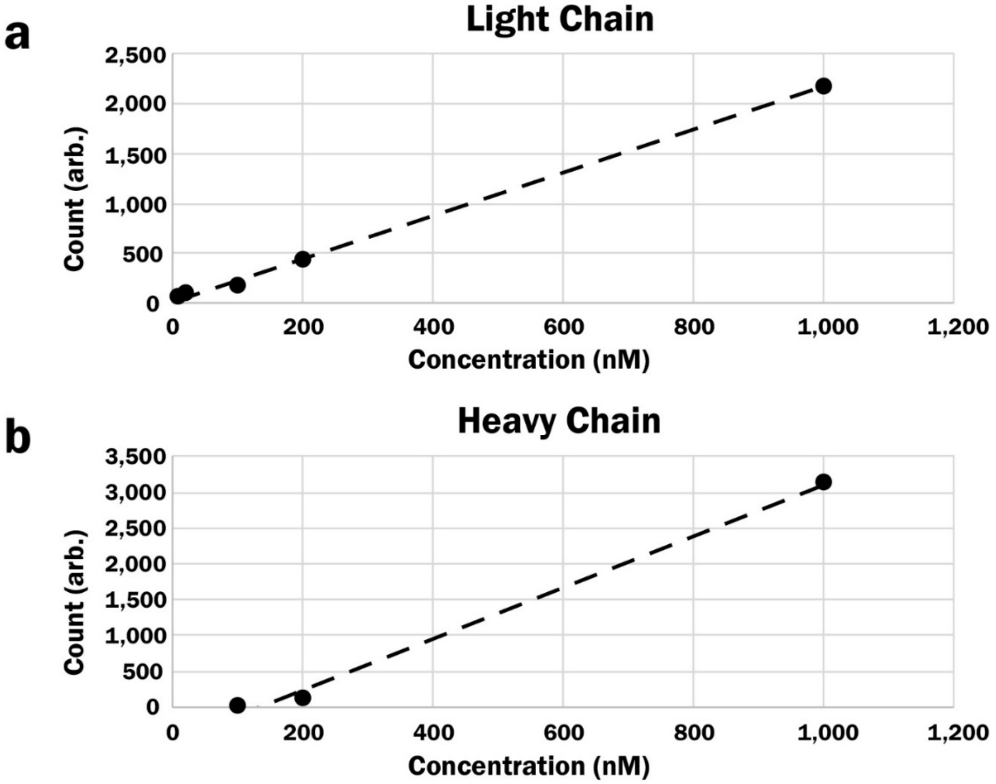


**Supplementary Fig. 4**. **Titration curve for Ig-MS to estimate the limit of detection (LOD) for NIST standard monoclonal antibody.** Standard curves for (a) light chain (LC) and (b) heavy chain (HC) with observed intensities and concentrations are shown. The LOD for the HC was ~100 nM, whereas that for the LC was not reached at the 10 nM data point; the LC LOD is estimated at <1 nM.

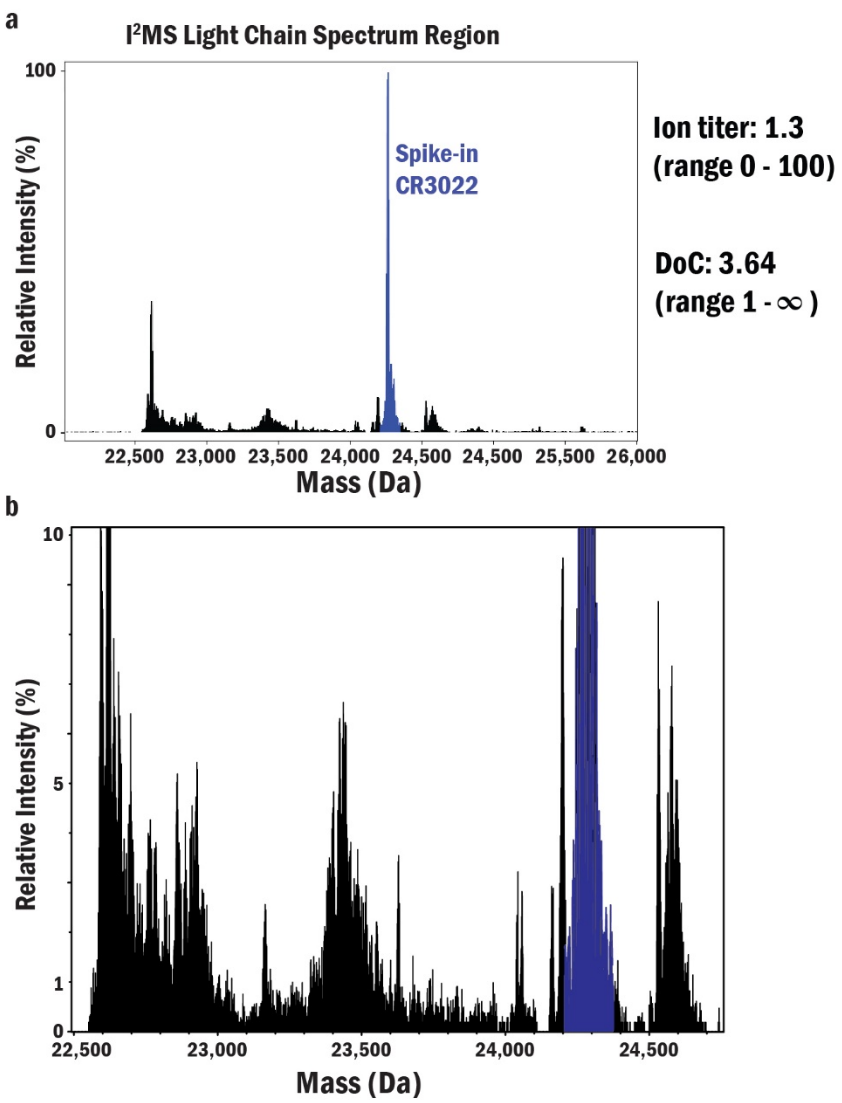


**Supplementary Fig. 5. Ig-MS readout of light chain region for CS1.** (a) Mass spectrum of all light chains isolated from plasma sample CS1. (b) Zoom in at 10% intensity to show LC peaks that correspond to 1% of the standard intensity. The peak highlighted in blue is the spiked-in mAb CR3022 at 100 ng.

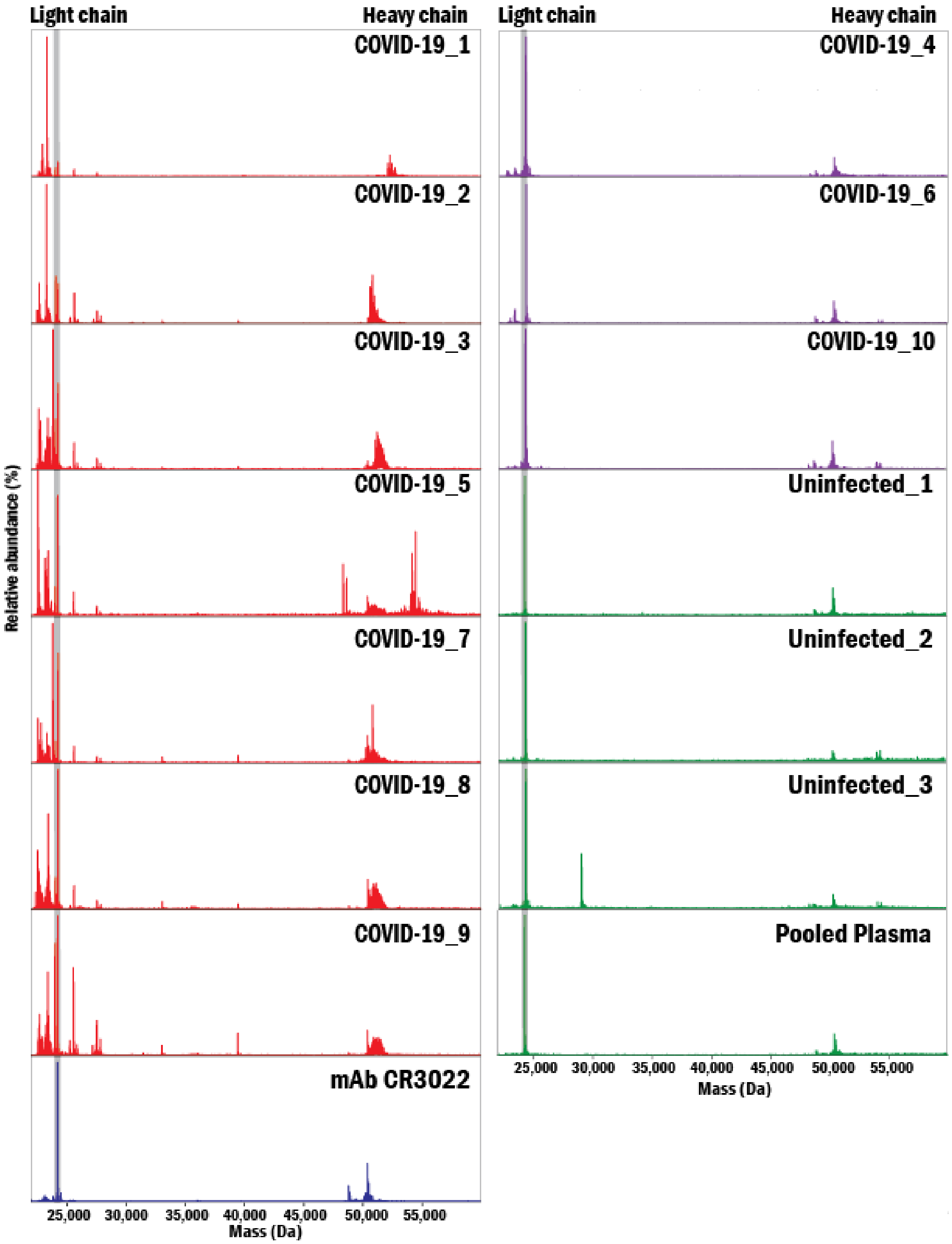


**Supplementary Fig. 6. Ig-MS readouts from Workflow 1 (Ig Light and Heavy Chains).** Results from plasma of ten COVID-19 convalescent patients, including seven hospitalized patients (red) and three outpatients (purple). Also, Ig-MS was applied to the plasma of three uninfected individuals, and a pool of plasma collected before the emergence of SARS-CoV-2 (green). Monoclonal antibody CR3022 anti-SARS-CoV-2-RBD (blue) was used as positive control and is highlighted in gray.

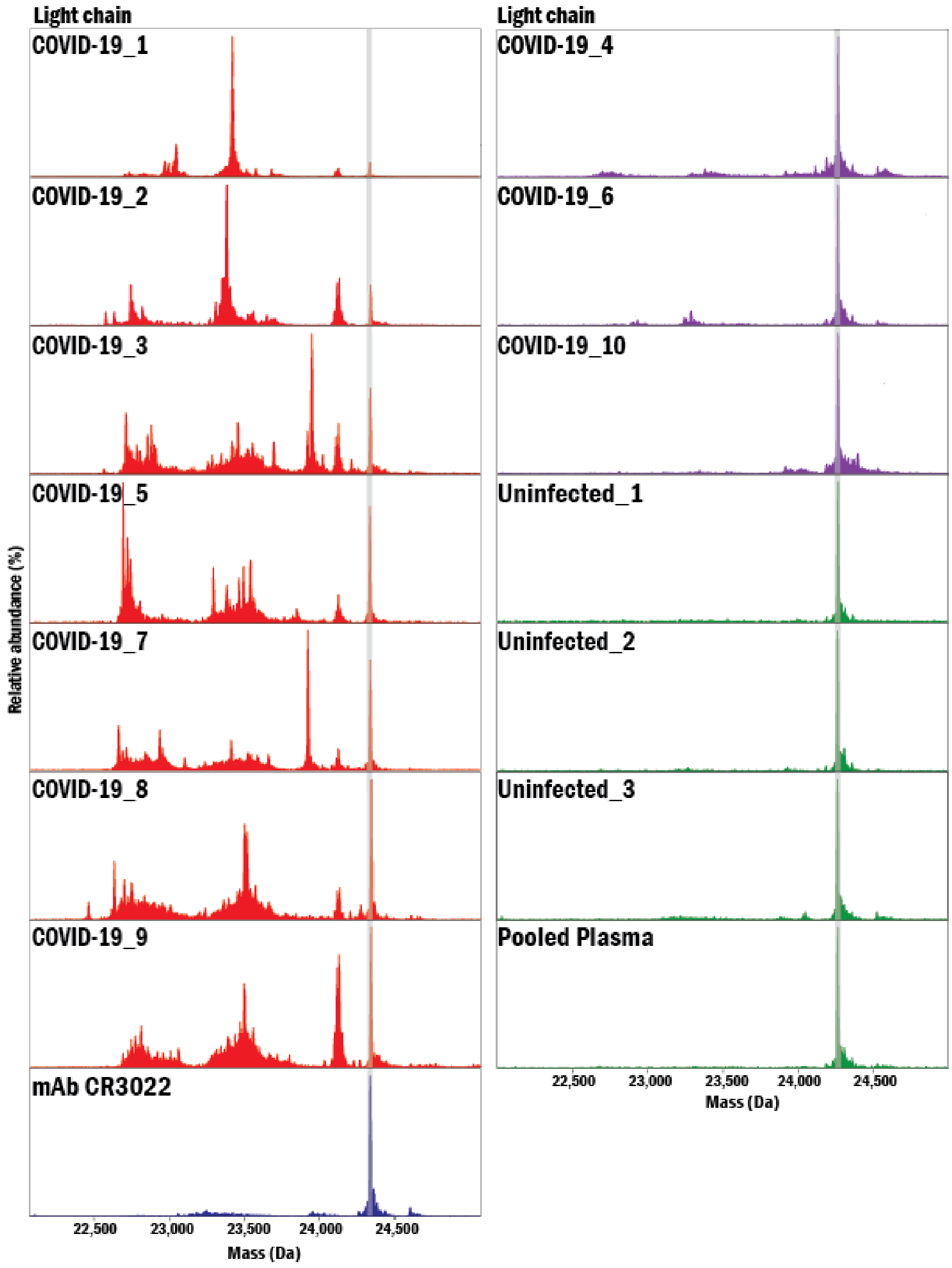


**Supplementary Fig. 7. Ig-MS readouts of Light Chains obtained using Workflow 1.** Plasma of ten COVID-19 convalescent patients, including seven hospitalized patients (red) and three outpatients (purple). In parallel, the assay was applied to the plasma of three uninfected individuals, and a pool of plasma collected before the emergence of SARS-CoV-2 (green). Monoclonal antibody CR3022 anti-SARS-CoV-2-RBD (blue), used as a positive control for quantitation, is highlighted in gray.

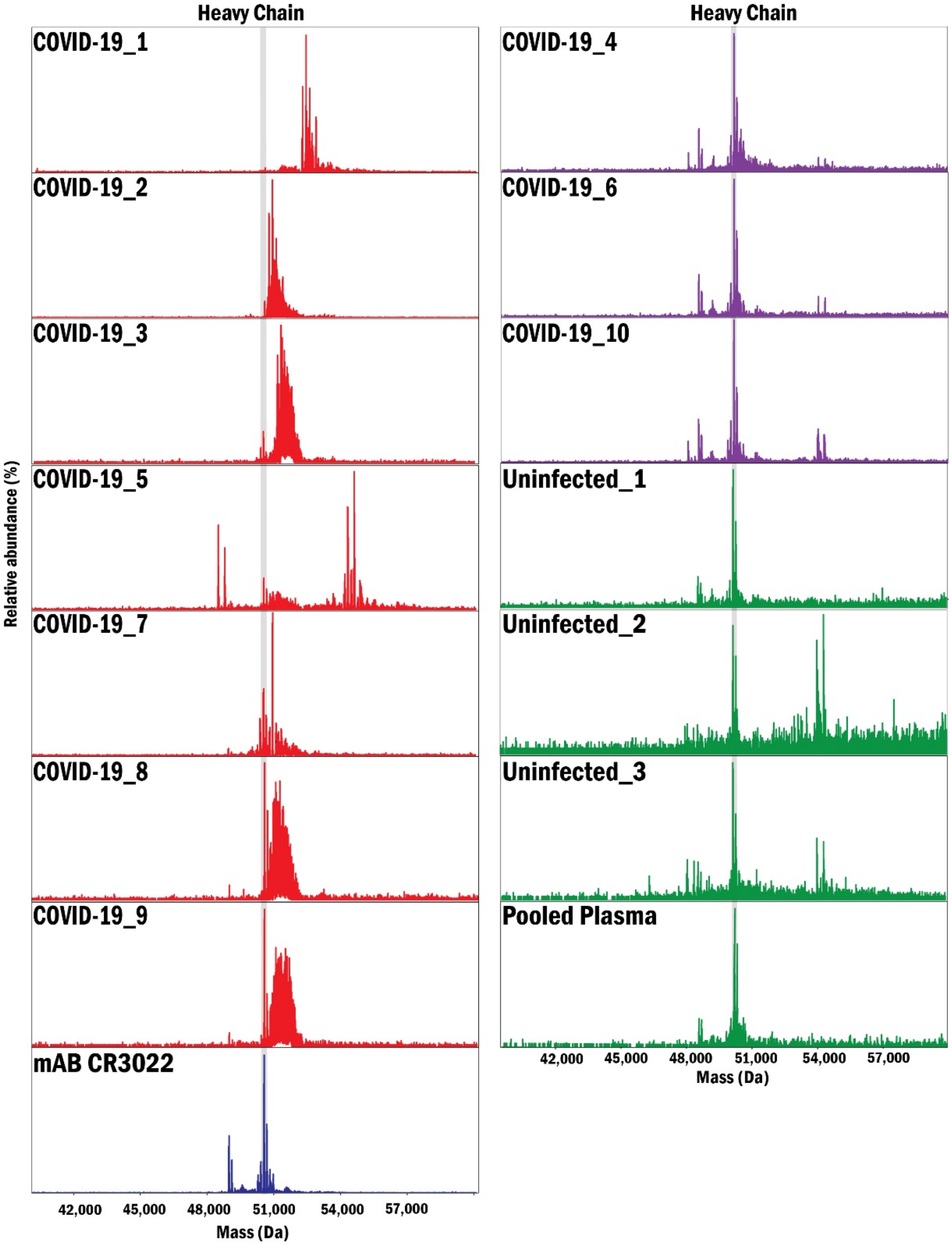


**Supplementary Fig. 8. Ig-MS readouts of Heavy Chains obtained using Workflow 1.** The plasma of ten COVID-19 convalescent patients, including seven hospitalized patients (red) and three outpatients (purple). Also, Ig-MS was applied to the plasma of three uninfected individuals, and a pool of plasma collected before the emergence of SARS-CoV-2 (green). Monoclonal antibody CR3022 anti-SARS-CoV-2-RBD (blue) spectrum was used as a positive control.

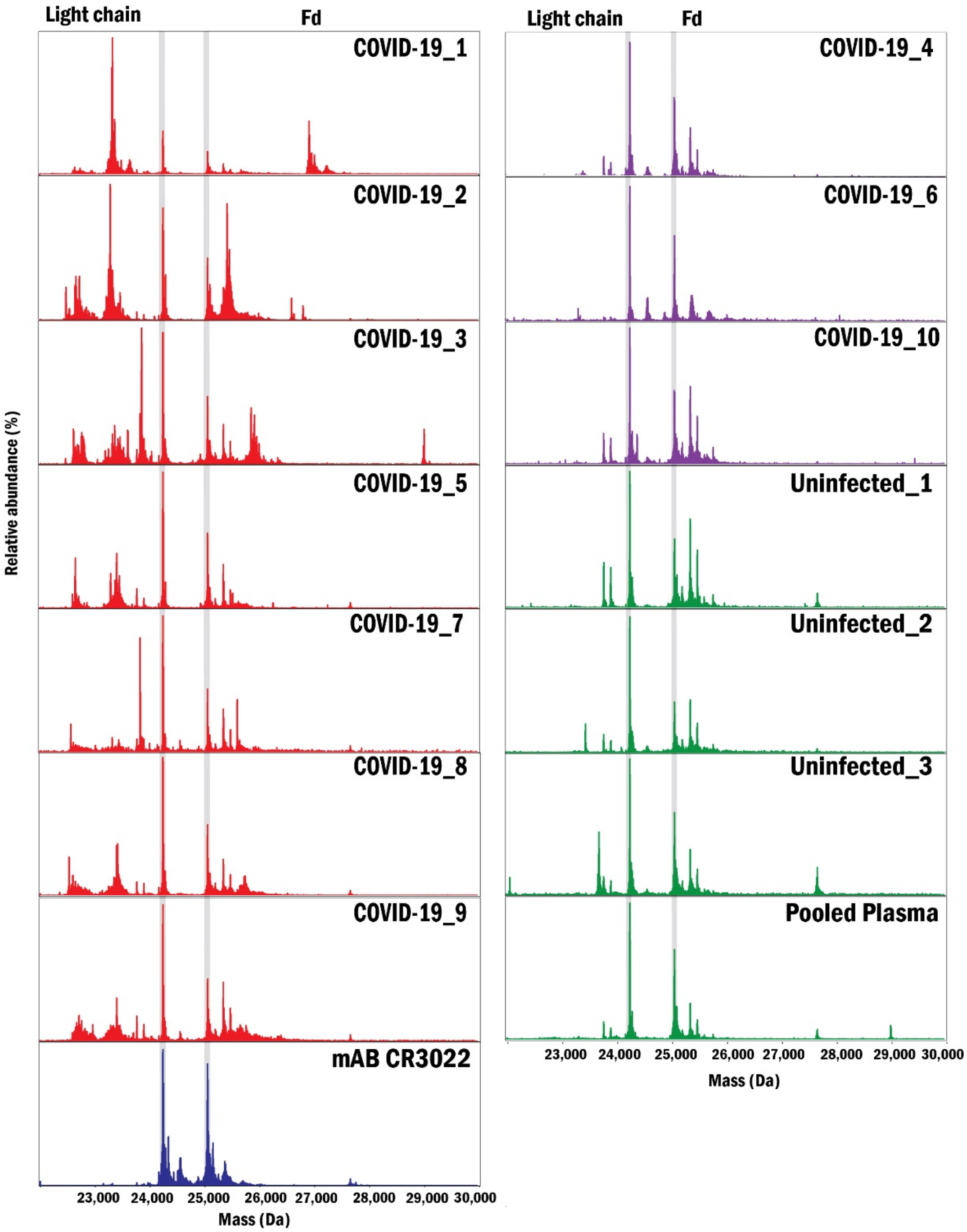


**Supplementary Fig. 9. Ig-MS results from Workflow 2 (light chain and Fd fragment from the heavy chain).** The plasma of ten COVID-19 convalescent patients, including seven hospitalized patients (red) and three outpatients (purple). In parallel, the assay was applied to the plasma of three uninfected individuals and a pool of plasma collected before the emergence of SARS-CoV-2 (green). Monoclonal antibody CR3022 anti-SARS-CoV-2-RBD (blue) was used as a positive control.

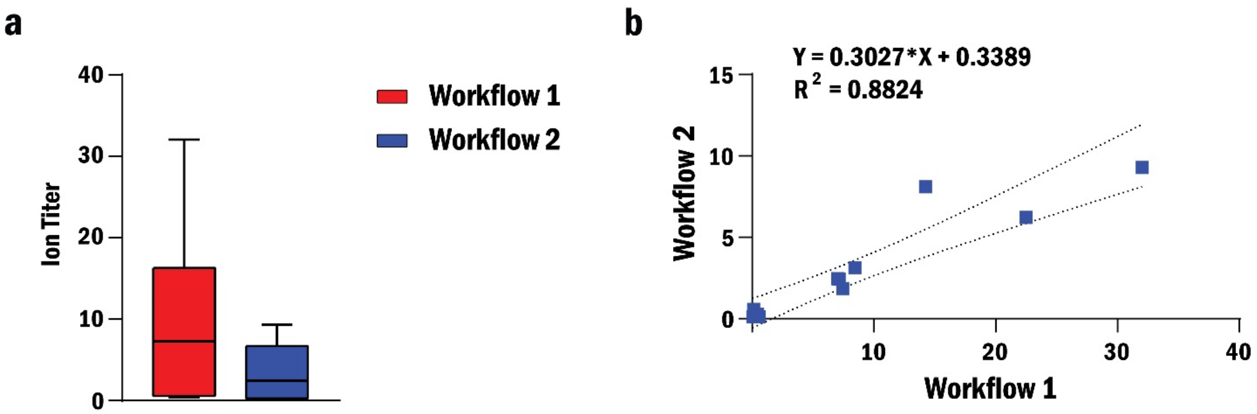


**Supplementary Fig. 10. Comparison of ion titers between Workflow 1 and Workflow 2.** (a) Box-and-whisker plot demonstrating the spread of ion titers obtained with the two Ig-MS workflows for light chain repertoires from ten COVID-19 patients. (b) Correlation between ion titers obtained with the two workflows.

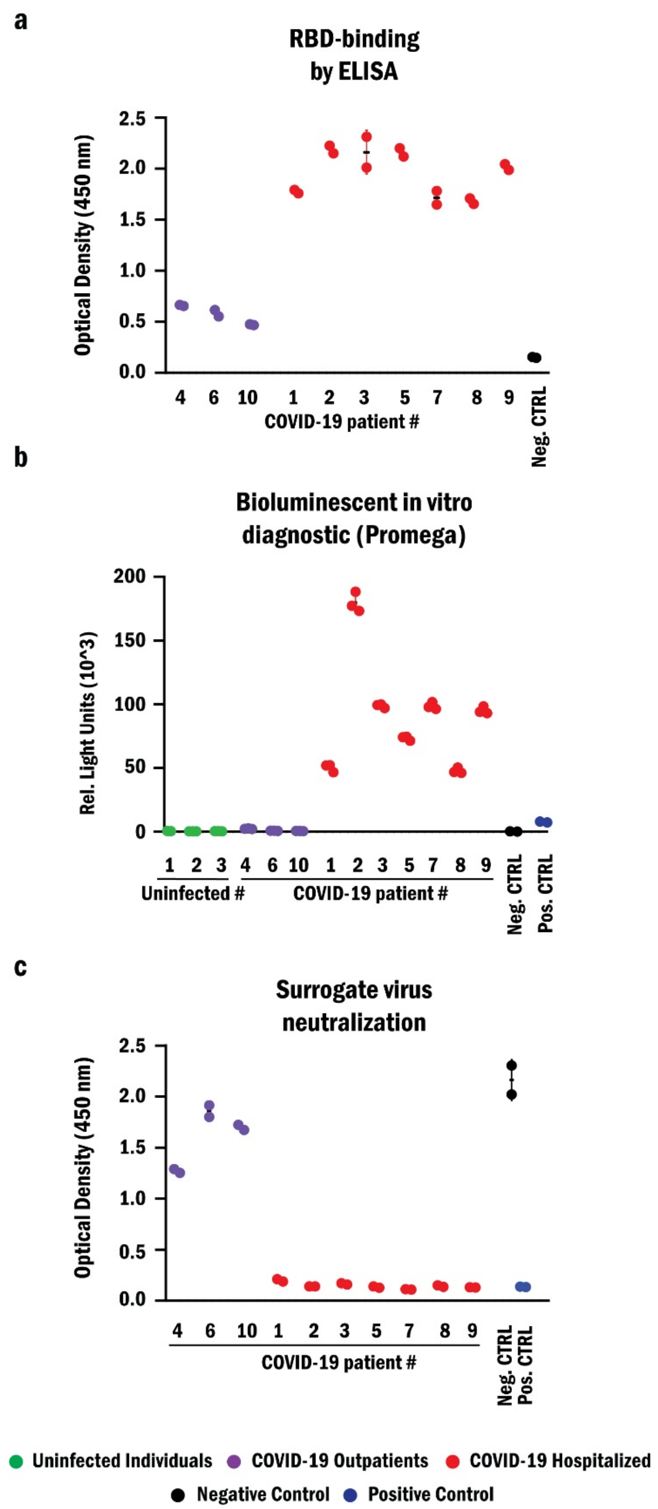


**Supplementary Fig. 11**. **Anti-RBD antibody and neutralization titers.** Titers and neutralization capacity of anti-RBD antibodies from COVID-19 outpatients (purple), hospitalized patients (red), and uninfected (green) calculated by (a) ELISA, (b) bioluminescent in vitro diagnostic, and (c) surrogate virus neutralization.

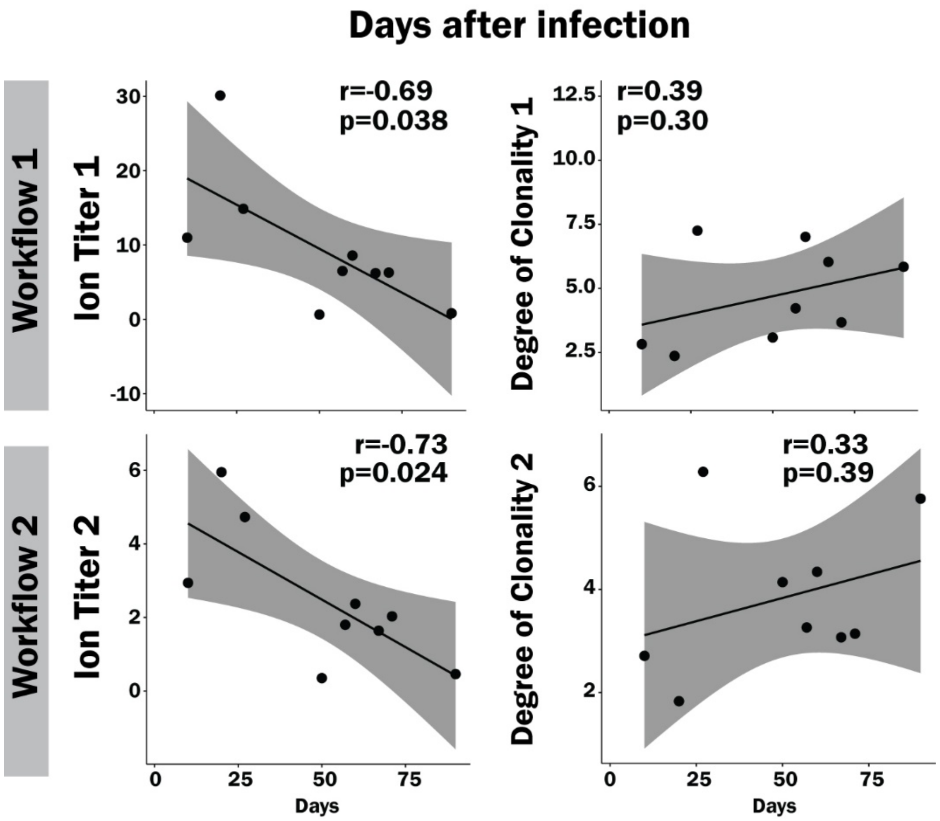


**Supplementary Fig. 12**. **Correlation of Ion titer (IT) and Degree of Clonality (DoC) metrics from Ig-MS with days after infection.** Shown are the Pearson correlation coefficient (r) of the Ion Titer and the Degree of Clonality from Workflows 1 and 2 against days after the infection. P-value (p).

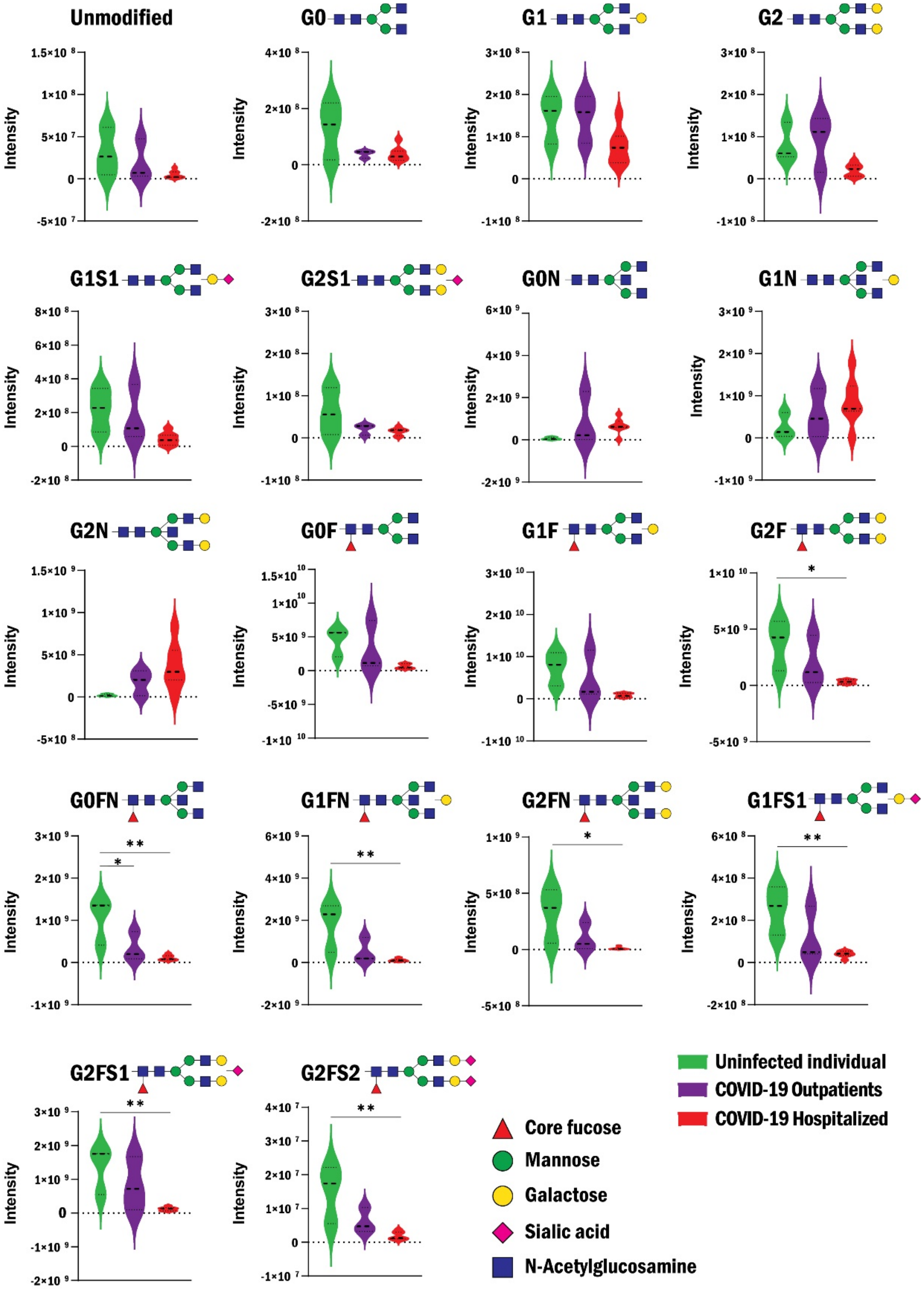


**Supplementary Fig. 13. Comparison of bulk IgG glycosylation profiles**. COVID-19 patients hospitalized (red) and outpatients (purple) with those of uninfected individuals (green). Statistical significance was calculated by one-way ANOVA with Tukey's multiple comparison test (*, p<0.05; **, p<0.01).

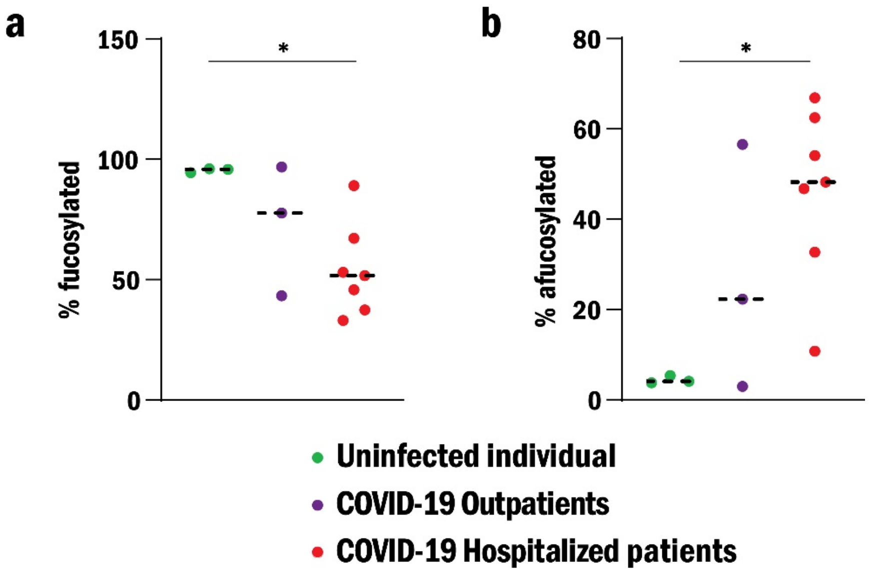


**Supplementary Fig. 14. Comparison of total glycan composition**. Percentage of total (a) fucosylated and (b) afucosylated glycans observed on COVID-19 patients hospitalized (red) and outpatients (purple) with those of uninfected individuals (green). Statistical significance was calculated by one-way ANOVA with Tukey's multiple comparison test (*, $p<0.05$)
